# Self-help digital psychosocial intervention for older adults with subthreshold depressive symptoms in primary care in Brazil (PRODIGITAL): Protocol for an individually randomised controlled trial

**DOI:** 10.1101/2023.04.06.23288257

**Authors:** Thiago Vinicius Nadaleto Didone, Carina Akemi Nakamura, Nadine Seward, Felipe Azevedo Moretti, Monica Souza dos Santos, Mariana Mendes de Sá Martins, Luara Aragoni Pereira, Evelyn da Silva Bitencourt, Marcelo Oliveira da Costa, Caio Hudson Queiroz de Souza, Gabriel Macias de Oliveira, Marcelo Machado, Jamie Murdoch, Pepijn Van de Ven, William Hollingworth, Tim J. Peters, Ricardo Araya, Marcia Scazufca

## Abstract

Subthreshold depression is a substantial risk factor for the development of major depression. It is associated with poorer health, functional disabilities, and reduced quality of life in older adults. There is a paucity of cost-effective psychosocial interventions for this population in primary care worldwide, particularly in low- and middle-income countries. We present a protocol for evaluating the effectiveness and cost-effectiveness of the Viva Vida programme, a 6-week self-help digital psychosocial intervention for treating older adults with subthreshold depressive symptoms in primary care with a two-arm, individually randomised controlled trial with a 1:1 allocation ratio with integrated economic and process evaluations. We will include 450 individuals 60 years and older with subthreshold depressive symptoms (score between five and below 10 on the 9-item Patient Health Questionnaire (PHQ-9)), registered with one of the 46 primary care clinics in Guarulhos, Brazil. The Viva Vida programme will be delivered via automated audio and visual WhatsApp messages, with psychoeducation and behavioural activation content. It involves 48 messages, delivered twice daily, four days a week. Participants in the control arm will receive a single message with general information about depression. The primary outcome is the depressive symptoms at the three-month follow-up as measured by PHQ-9, which will be compared between study arms. The cost-effectiveness of the intervention will be assessed at five months. A detailed process evaluation will explore context and implementation outcomes. The Viva Vida programme is an innovative digital psychosocial intervention delivered via WhatsApp messaging without the participation of health professionals. The evaluation of the Viva Vida programme will contribute to the development of simple and cost-effective models of remote self-help interventions for reducing depressive symptoms among older adults with subthreshold depression in primary care. The protocol is registered with Registro Brasileiro de Ensaios Clinicos (ReBEC) (RBR-6c7ghfd).

## Introduction

The development of simple and effective treatments for older adults with subthreshold depression is an important public health issue and has been identified as a neglected area of research [1–3]. The definition and diagnostic criteria of subthreshold depression (also known as subsyndromal depression, subclinical depression or minor depression) are heterogeneous [1]. Broadly speaking, subthreshold depression requires the presence of at least one core symptom of depression (depressed mood or anhedonia), while the severity of symptoms does not meet the criteria for a diagnosis of depression [4,5].

The negative impact of subthreshold depression on the health of older people and healthcare systems is well known. Subthreshold depression is associated with functional disability, reduced quality of life, loneliness, suicide attempts, mortality and increased healthcare utilisation [1,6–8]. Furthermore, the high prevalence of subthreshold depression among older adults in the community, the persistence of symptoms over time, and the progression to moderate and severe forms of depression in many cases highlight the public health challenge of addressing this condition [3].

A review of trials conducted in the 2000s to reduce depressive symptoms in older, community-dwelling adults with subthreshold depression recommended the use of psychotherapy [9]. This review showed that psychotherapeutic approaches (i.e. cognitive behavioural therapy, problem solving therapy, physical activation and behavioural activation) were safe and effective in reducing depressive symptoms in primary care settings [10–13]. However, only one study in this review evaluated the effectiveness of a remote intervention method using a web-based self-administered cognitive behavioural therapy [11]. More recently, the UK Casper trial provided evidence of the effectiveness of a collaborative care programme, delivered mainly remotely, in reducing depressive symptoms and progression to moderate and severe symptoms in older adults with subthreshold depression in primary care [14]. The Casper programme has innovative components. Nursing assistants delivered a rapid and relatively inexpensive behavioural activation programme over the telephone, improving access to treatment.

The availability of remote mental health interventions has recently increased [15], driven by the COVID-19 pandemic. A recent umbrella review of 38 systematic reviews showed that the restrictions on social contact between people imposed by COVID-19 measures have impacted how mental health services are provided. Synchronous tools (e.g. video conferencing) and, to a lesser extent, asynchronous tools (e.g. mobile applications) have been used to provide remote mental health care to patients, particularly in high-income countries [16]. However, the scarcity of evidence of research in primary care settings worldwide does not allow us to determine which forms of synchronous or asynchronous self-help telepsychosocial approaches are effective for older adults with subthreshold depression. Identifying cost-effective psychosocial treatments and the older adults who are most likely to benefit from them is urgent in low- and middle-income countries (LMICs), where primary care is struggling to meet the physical and mental health needs of a rapidly growing older population.

We will conduct a randomised controlled trial to evaluate the effectiveness and cost-effectiveness of the Viva Vida programme (PRODIGITAL), a 6-week digital psychosocial intervention delivered via automated audio and visual WhatsApp messages for the treatment of older adults with subthreshold depressive symptoms in primary care in Guarulhos, Brazil. A process evaluation using qualitative methodology will examine the implementation outcomes (acceptability, appropriateness, fidelity, and feasibility) of the intervention, the contextual barriers and facilitators to implementing this programme.

## Methods

We present the protocol of a two-arm (1:1 allocation ratio) randomised controlled trial (RCT) with integrated economic and process evaluations.

### Study setting

The study will recruit individuals registered at 46 primary care clinics in underprivileged areas of Guarulhos, known as Unidades Básicas de Saúde (UBSs). All primary care clinics have Family Health Teams (FHTs) and care is based on the Family Health Strategy model. Guarulhos has a population of approximately 1.4 million. The average per capita household income is BRL 791 reais (approximately USD 159 dollars in 2023) [17]. In Guarulhos, 13.8% of the population is 60 years or older, of which 55.7% are women [18], and 14.6% are illiterate [19]. In the Southeast region of Brazil, where Guarulhos is located, 75.7% of older adults have a mobile phone [20] and 64.9% have connected to the Internet in the last three months [21].

### Participants

#### Inclusion criteria

a. Individuals aged 60 years or over registered with one of the 46 participating UBSs;
b. Individuals able to receive and listen to WhatsApp messages;
c. Individuals screening positive for subthreshold depressive symptoms assessed with the 9-item Patient Health Questionnaire (PHQ-9), defined as a score of at least 1 on the PHQ-2, which includes the first two questions of the PHQ-9 (depressed mood and anhedonia), and a PHQ-9 total score of at least five and less than 10 [4,22,23].

#### Exclusion criteria

a. Individuals with communication problems (e.g., non-Portuguese speaking or cognitive impairment to the extent that these interfere with study assessments or to participate in the intervention);
b. Individuals with visual or hearing impairments to the extent that they interfere with study assessments or to participate in the intervention;
c. Individuals who are unable to participate in the study for five months;
d. Individuals identified as being at acute suicide risk (i.e. a suicide attempt in the two weeks before the screening assessment) using the Immediate Suicide Risk Protocol [24];
e. Individuals living in the same household as another study participant;
f. Individuals who participated in the PROACTIVE trial [25].

### Interventions

Participants in the intervention arm will receive a self-help digital psychosocial intervention delivered via audio and visual WhatsApp messages. The control arm will receive a single audio message. The research team will not interfere with the health care that participants may receive during and after their participation in the trial.

#### Digital psychosocial intervention

The Viva Vida programme was adapted from the short, animated videos shown by community health workers to older adults with depressive symptomatology in the RCT testing the PROACTIVE intervention [24,25] and grounded in the theories of psychoeducation [26] and behavioural activation [27]. Behavioural activation is an approach in mental healthcare that aims to promote behaviours that activate positive emotions, as well as avoid those that worsen depressive symptoms, and integrate them into one’s routine [27]. The programme consists of 48 messages that will be sent to participants via WhatsApp over six weeks, four days per week. Each day, they will receive one message in the morning and one in the afternoon. No support of health professionals will be provided as part of the intervention.

There are two types of messages: audio messages and images with short texts. Audio messages last, on average, three minutes and are based on the storytelling technique, a communication tool that can attract attention, arouse emotions and engage listeners to influence their attitudes and beliefs [28]. We chose audio messages because of the low level of education and income of the older adults living in disadvantaged areas of Guarulhos. The audio messages are short to make Viva Vida viable for older people with mobile phones with low storage capacity. Through the audio messages, two fictitious characters (Mrs. Zuzu and Mr. Zé) share experiences as if they had participated in the Viva Vida programme. These characters share what they have learned about depression (psychoeducation), how they have felt, and what kind of pleasant and meaningful activities they have done, and the ones they have avoided to increase positive interactions with people and the environment (behavioural activation). The messages also include health promotion tips, such as information on diet, physical activity, the importance of other health treatments, and relapse prevention. The audio and visual messages advise participants to contact health services if they feel that their depressive symptoms are not improving or are getting worse. The characters use language, vocabulary, tone and manner appropriate to the target audience. They are realistic to create empathy for the study participants when listening to the messages. The content of the Viva Vida programme follows with the WHO recommendations for digital interventions for depression [29] and the theoretical basis of interactive health communication applications [30,31]. It also follows guidelines suggesting that patients should be educated about depression and encouraged to self-manage their symptoms [32,33].

To show how behavioural activation technique is embedded into the Viva Vida programme, we present an example of one of the audio messages in which Mrs. Zuzu practises what she has learned during the Viva Vida programme. She shares with the listener her experience of doing an activity she enjoys has helped to improve her mood.

> *(Mrs. Zuzu, message 11, week 2) “I want to tell you something that happened to me the other day. I woke up upset, feeling down, and I thought to myself I have to get up and do something. I have a little garden here at home and I love to plant and take care of the plants. At first, I felt discouraged, but there I was, watering the lettuce, and working the soil. You know, after a while I got so distracted that I didn’t even realise I was feeling discouraged. It was really cool. That was when I realised that doing something we like, something we care about, changes our mood. It works. And if it works for me, it will work for you too. Give it a try!”*

As part of the programme, participants will receive one extra message at the end of each week, using the WhatsApp quick reply tool, with questions about their experience of participating in Viva Vida. These questions will investigate whether the messages helped them understand the signs of depression, if they could choose and do activities that may help them feel better, whether the Viva Vida programme is helping them to feel better, and whether they liked being part of the programme. The WhatsApp quick reply tool allows participants to answer the question by selecting one pre-defined answer, for example, *yes*, *more or less*, or *no*. After replying, participants will receive an automated pre-recorded audio response. These messages will also invite participants to share further comments about the programme by sending spontaneous WhatsApp messages.

#### Single message

Participants allocated to the control arm will receive a single audio message of approximately six minutes via WhatsApp. This message informs them about the main signs of depression, simple ways to improve their mood, and advises them to contact health professionals for further support if they do not feel better or need additional care.

### System to deliver the messages

The technical research team will coordinate the delivery of the messages. We will use a web system integrated with the WhatsApp Business Application Programming Interface (API), which is managed by an intermediary company. This system is hosted on a cloud service where access is restricted by both authentication and authorisation processes. Data flowing to and from browsers is also protected through the use of encryption. The WhatsApp Business API allows the research team to capture the date and time (timestamp) when messages are sent, delivered and opened, replies to the quick reply tool, and the content of spontaneous messages. The API enables customised messages (e.g. messages with the recipient’s name) and the attachment of audio and image files. Messages will be scheduled weekly and delivered using a job scheduler on the server.

During the first two weeks of the programme, a member of the technical support team (who does not take part in the other activities of the study) will contact participants who are not receiving or opening the messages to check if they are experiencing any technical problem. We will identify these participants through the Viva Vida web dashboard, which will show the status of messages as *sent to intermediary company platform*, *sent to participant*, *delivered to participant*, or *opened by the participant*. During the remaining four weeks of the Viva Vida programme, participants will be able to contact the technical support team if they are not receiving the messages.

We will remove participants who block their mobile phones from receiving the Viva Vida messages to comply with the WhatsApp policy and keep our system functional. These participants will not receive new intervention messages, but will be contacted again during follow-up evaluations unless they contact us to withdraw their consent to participate in the study.

### Outcomes

Follow-up assessments will take place three and five months after sending the first message (intervention arm) or the single message (control arm).

#### Primary outcome

The primary outcome will be the (continuous) PHQ-9 score as a measure of depressive symptoms at the three-month follow-up, which will be compared between study arms as allocated.

#### Secondary outcomes

Depressive symptoms(mean PHQ-9 score) at five months. Other secondary measures assessed at three and five months will be the proportion of participants with clinically significant depressive symptomatology (PHQ-9≥10), change in anxiety symptomatology as measured by the 7-item Generalised Anxiety Disorder (GAD-7) [34], loneliness as measured by the 3-item University of California, Los Angeles (UCLA) loneliness scale (3-item UCLA) [35], quality of life measured by the European Quality of Life five-dimensional questionnaire, five-level version (EQ-5D-5L) [36], and capability well-being measured by the ICEpop CAPability measure for Older people (ICECAP-O) [37]. The cost-effectiveness of the Viva Vida programme will be assessed at the five-month follow-up.

### Sample size

To detect what we consider to be a relevant clinical difference between the two randomised arms of 0.33 standard deviations [14], 142 to 162 individuals in each arm gives 80% to 85% power at a two-sided 5% significance level. Data from our PROACTIVE pilot study [38] (PHQ-9 means of 5.9 and 6.4 with a standard deviation of 1.5; data not published) suggest that this effect size is feasible for this kind of intervention. We anticipate 25% attrition and therefore plan to recruit 225 individuals in each arm, for a total sample size of 450.

### Recruitment

The Guarulhos Health Secretariat will provide a list with contact details of all individuals aged 59 years or older who are registered in the participating UBSs (the slightly lower age limit compared to the relevant inclusion criterion is so that we include individuals who would be 60 years old during the recruitment period). Names duplicated on the list, individuals without a mobile phone number, and participants from our previous RCT conducted in Guarulhos [24] will be excluded. The remaining individuals (alphabetical ordered) on the list will receive a random ID number. The recruitment will follow the ID number in ascending order. This new list will be used to simultaneously recruit participants for two RCTs evaluating a digital intervention for older adults with depressive symptoms being conducted by our research group [39]. These two studies will use the same protocol to recruit participants; only the inclusion criterion for the severity of depressive symptoms will be different. In the present study, we will recruit older adults with subthreshold symptoms of depression as defined above.

The research team will contact via WhatsApp message individuals who have at least one mobile phone number. This message will briefly describe the study and inform these individuals that a research assistant will contact them by phone. Only those whose WhatsApp message is successfully delivered (i.e. an active WhatsApp number) will be contacted by phone and invited to start the recruitment interview. This interview will consist of three stages:

a. Screening assessment: will assess inclusion criteria (age, WhatsApp use and assessment of depressive symptoms) and exclusion criteria (communication and engagement problems, acute suicide risk, and health conditions that prevent participation in the trial);
b. Baseline assessment: participants screened positive for subthreshold depressive symptoms and with no exclusion criteria will be assessed for anxiety, loneliness, quality of life, capability well-being, socioeconomic profile (marital status, race, education level, living arrangement, and personal and household income), and alcohol and tobacco use;
c. Invitation to participate in the study.

Whenever possible, the screening and baseline assessments will be carried out consecutively during the same telephone call. Eligible participants will then be invited to participate in the study. If the participant is unable to complete all three stages of the interview in the same phone call, the research assistants will arrange another time to complete recruitment no more than 28 days after the PHQ-9 screening. Before screening and inviting individuals to participate in the study, we will inform them about the study and obtain their consent. The full study timeline is shown in Fig 1, and a detailed diagram of the study procedures is shown in Fig 2.

**Fig 1.**
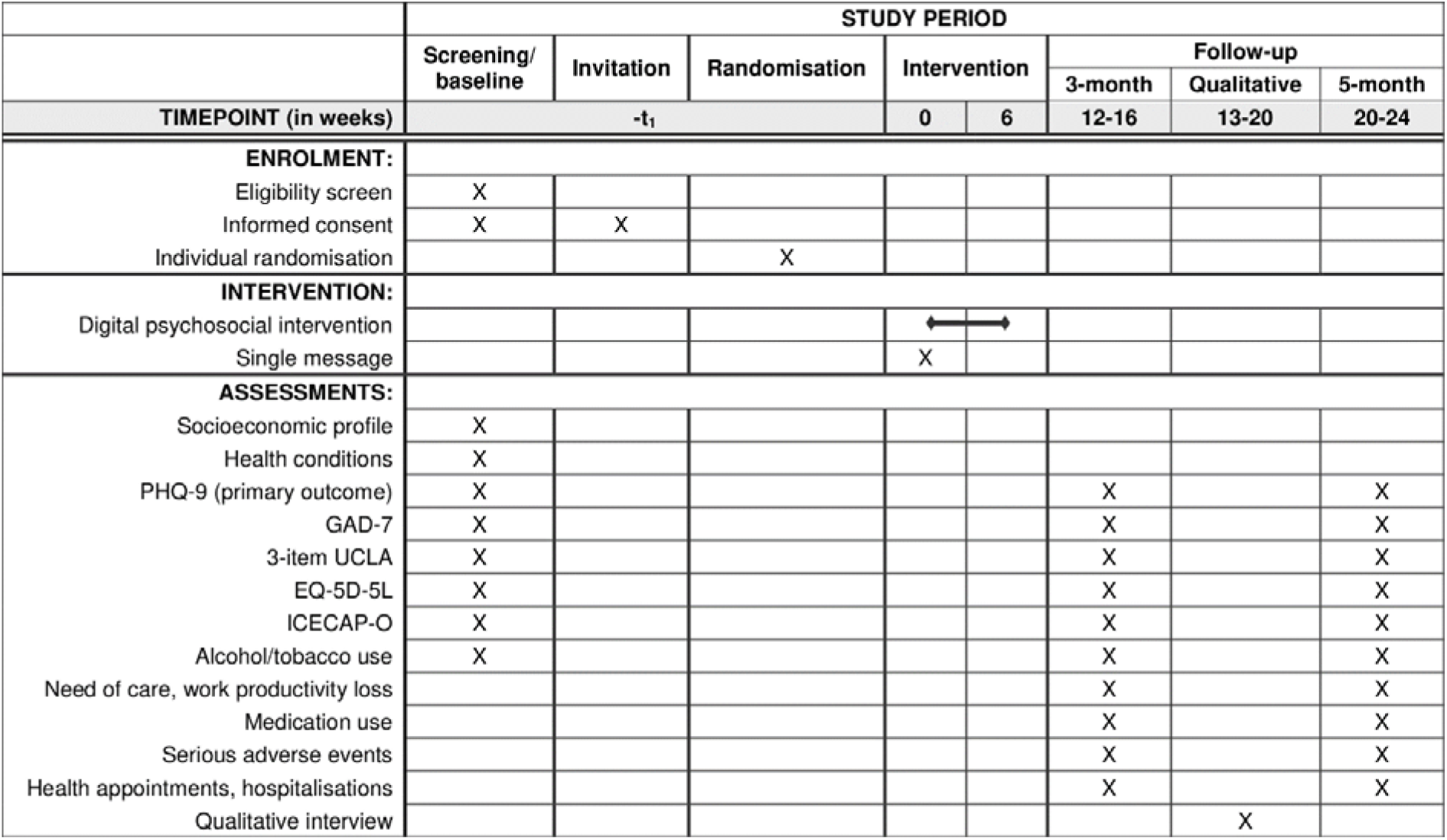
SPIRIT schedule of enrolment, interventions and assessments. 3-item UCLA, 3-item University of California, Los Angeles (UCLA) loneliness scale; EQ-5D-5L, European Quality of Life five-dimensional questionnaire, five-level version; GAD-7, 7-item Generalised Anxiety Disorder; ICECAP-O, ICEpop CAPability measure for Older people; PHQ-9, 9-item Patient Health Questionnaire. The qualitative interview will be conducted with 24 purposely selected participants from the intervention arm.

**Fig 2.**
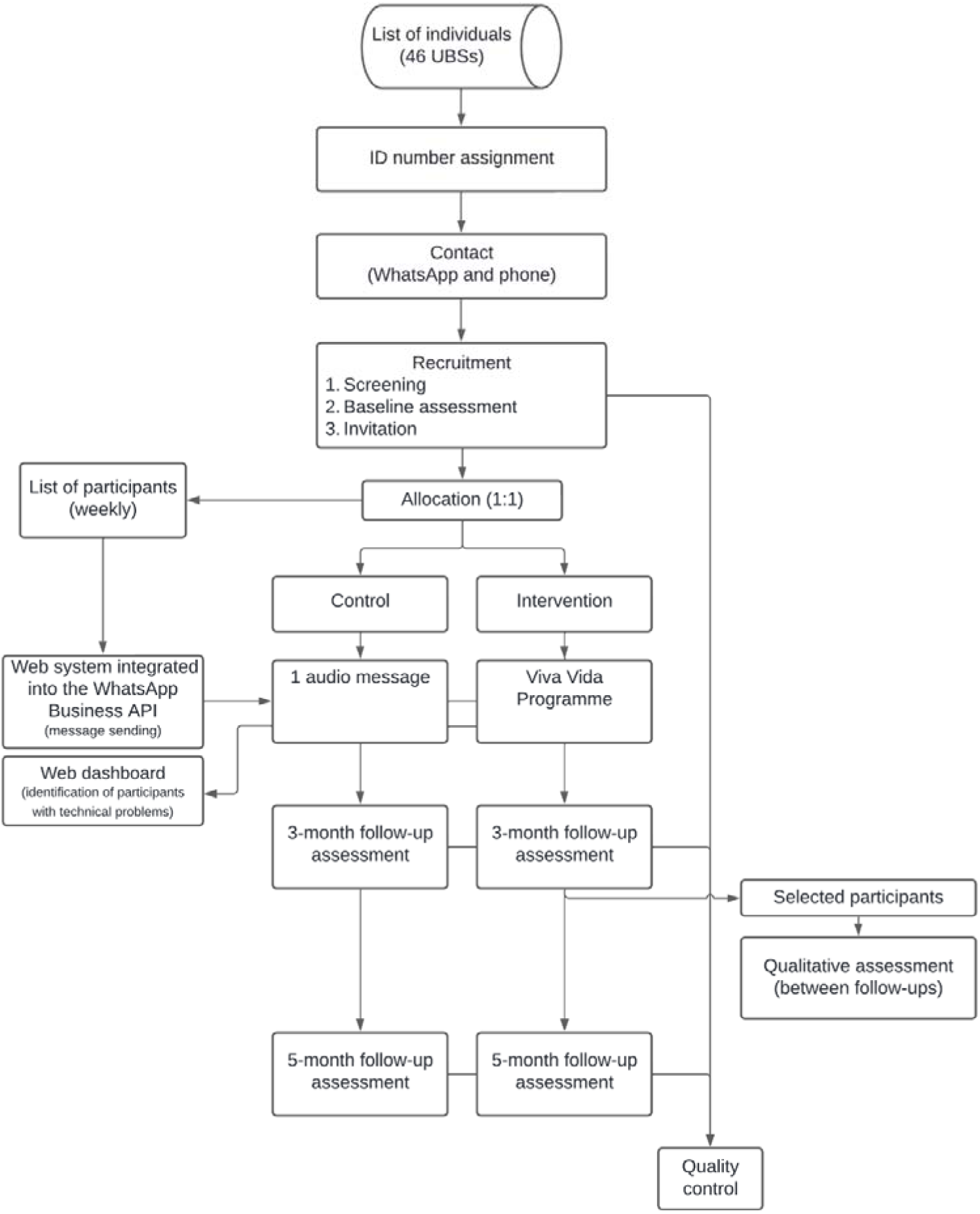
Diagram of the main procedures of the trial. Trained research assistants will contact individuals registered in 46 UBSs from Guarulhos, São Paulo, Brazil. After a three-step recruitment interview, participants will be allocated (1:1) to control (one audio message) or intervention (Viva Vida programme) arms. A web system will send messages to both arms, and a web dashboard will identify technical problems. Follow-up assessments at three and five months will be conducted. Qualitative assessments will be made in selected participants between follow-up assessments. Quality control will be performed in a sample of baseline and follow-up assessments. API, Application Programming Interface; UBS, primary care clinics known as Unidades Básicas de Saúde.

### Allocation

Once individuals have been recruited, they will be randomly allocated in a 1:1 ratio to either the intervention or control arm and placed on a list to receive their allocated intervention, which will be sent out within a 10-day window. Therefore, each list will contain participants recruited in the previous week.

Two members of the research team not directly involved in the recruitment (CAN and TJP) will generate the randomisation allocation sequence using Microsoft Excel, using random permuted blocks with random block sizes. Stratification will be based on gender (women/men), age groups (60-69/70-79/≥80 years) and type of primary care clinic model (Family Health Strategy or mixed model – Family Health Strategy and traditional primary care). Only participants seen by Family Health Strategy teams will be included in the mixed model clinics. The allocation sequence will be concealed in the randomisation module of the Research Electronic Data Capture (REDCap) software [40,41].

### Blinding

Given the differences in the number and content of the messages received by the intervention and control groups, blinding of participants will not be feasible. The team of researchers responsible for recruitment and follow-up assessments and the team responsible for the intervention will work independently. Research assistants conducting recruitment and follow-up assessments will be blinded to group allocation. Whenever possible, the same research assistant will not conduct more than one interview (recruitment or follow-up) with the same participant. The risk of contamination is therefore very low. Messages sent to participants in both arms of the study will be delivered remotely, and participants will not be informed about other people enrolled in the RCT.

### Data collection and management

Data will be collected and managed using the REDCap software, hosted at the Hospital das Clínicas da Faculdade de Medicina da Universidade de Sao Paulo [40,41]. REDCap is a secure, web-based software platform designed to support data capture for research studies, providing: 1) an intuitive interface for validated data capture; 2) audit trails for tracking data manipulation and export procedures; 3) automated export procedures for seamless data downloads to standard statistical packages; and 4) procedures for data integration and interoperability with external sources. Recruitment and follow-up will be conducted by telephone by independent research assistants. Research assistants will be trained in the correct use of research questionnaires and data entry into the REDCap software. They will obtain permission from participants to record the interviews. The recorded interviews will be used to assess the quality of data collection.

Assessments of depressive symptomatology (PHQ-9), anxiety symptomatology (GAD-7), loneliness (3-item UCLA), health-related quality of life (EQ-5D-5L), capability well-being (ICECAP-O), alcohol and tobacco use will be administered at recruitment and repeated at three- and five-month follow-up. We will also ask participants if they need help with activities of daily living, recent time lost from work, consultations with health professionals (including mental health professionals), hospital admissions, and ongoing treatment for depression during the follow-up period. Severe adverse events associated with trial participation, such as suicide attempts, hospitalisation and death will be assessed for all participants in both follow-up assessments. At the end of both follow-up assessments, participants allocated to the intervention arm will be asked about their experience with the Viva Vida programme. We will collect data from the electronic health system’s records on psychotropic medication use and consultations with general practitioners and nurses for all participants during the five months they were enrolled in the RCT. This will be done shortly after the end of the last follow-up.

An independent researcher, who will not conduct interviews with participants, will carry out quality control of the recruitment and follow-up assessments. A sample of the recorded interviews will be reviewed to ensure adherence to the script and the quality of data collected.

We will leave a 4-week window for the follow-up assessments to minimise attrition. The three-month follow-up will take place between the 12^th^ and 16^th^ week, and the five-month follow-up between the 20^th^ and 24^th^ week after sending the first message (intervention arm) or the single messages (control arm). If the research assistants have difficulty contacting participants by phone during the follow-up window, then they will send an audio and/or a text message via WhatsApp to try to remind participants of the interview. If there is no response, the UBSs managers will be contacted to assist in reaching the participant. As a last resort, after three weeks of unanswered contact attempts, a research assistant will visit the participant at home to conduct a face-to-face assessment.

### Statistical methods

The analysis of the primary (depressive symptoms at three months) and secondary (depressive symptoms at five months, proportion of clinically significant depressive symptomatology - PHQ-9≥10, and anxiety, loneliness, quality of life and capability well-being at three and five months) outcomes will follow the intention-to-treat (ITT) principle, i.e. participants will be analysed in the arm to which they were allocated (regardless of adherence to the intervention), and will follow the Consolidated Standards of Reporting Trials (CONSORT) guidelines for randomised trials [42]. No interim analyses are planned. Descriptive statistics will compare differences in baseline demographics and clinical characteristics between the intervention and control arms to identify any imbalances. Linear regression models adjusted for stratification and the relevant baseline score will be used to examine the primary and secondary continuous outcomes. For the binary outcome, logistic regression and Poisson regression models will be used to generate relative risks. For secondary analyses, models will include any baseline variables that are not balanced between arms.

Exploratory subgroup analyses will include the Wald test for interaction terms between the relevant baseline variable and trial allocation in the above regression models to examine whether baseline PHQ-9 scores, gender, age, education level, and comorbid physical illness (diabetes, hypertension, and both) modify the effect of the intervention on PHQ-9 scores at both follow-ups. The results of these analyses will need to be interpreted with caution due to the limited power to detect such interactions and the paucity of evidence on the theoretical basis for these hypotheses.

Additional analyses will include a Complier Average Causal Effect (CACE) analysis [43] using an instrumental variable estimator and imputed data to determine the effect of the number of messages listened to, on the reduction in depressive symptom severity at both three and five months. At the three-month follow-up, participants randomised to the intervention arm will be asked at the end of the assessment how many messages they listened to from start to finish, with the following options: *none*, *a few*, *at least half*, *most* or *all*. The CACE analysis will consider listening to at least *most* of the messages received as the threshold, as we hypothesise that this is the minimum number of messages that participants need to listen to in order to have a therapeutic effect. Alternative analyses will be conducted using the thresholds of (a) listening to at least half of the messages and (b) listening to all of the messages. We will also consider using the number of messages opened (as recorded by the system) as a continuous variable (if assumptions of linear relationships are met).

If we find any marked differences in missing data between the intervention and control arms, or a proportion of missing data greater than 10%, then we will impute missing data separately for each arm for all analyses described above. We will use multiple imputations by chained equations (MICE models), assuming data are missing at random (MAR) [44]. MICE models will include variables from the original analyses (outcomes, stratification and gender) and any other variables that predict missingness [45,46]. The selection model approach will be used to conduct sensitivity analyses testing for modest departures from the MAR assumption for primary outcomes only [47–49].

The cost-effectiveness analysis will compare the costs and effects of the Viva Vida programme against a single message from the health system’s perspective. The first analysis will estimate the incremental cost-effectiveness using (a) the primary clinical outcome measure (cost per patient recovered), and (b) the quality adjusted life years (QALYs) calculated using the EQ-5D-5L [36,50]. Incremental cost-effectiveness ratios and cost-effectiveness acceptability curves will be presented to show the probability of the intervention being cost-effective at a range of willingness-to-pay thresholds [36,50]. The second analysis will be the Net Monetary Benefit (NMB) statistic; it will be calculated at the WHO recommended threshold for LMICs using the difference in costs and the difference in QALYs between the two arms. The EQ-5D-5L responses will be converted to utility scores using the most appropriate population tariff for the Brazilian population available at the time of the analysis. QALYs will be estimated using utility scores adjusted for baseline values. We will use national, where available, or local unit costs to value resource use.

### Process evaluation and analysis

Qualitative interviews will be conducted to gain insight into participants’ reasons, motivations, modes and contextual barriers and enablers that may affect the clinical and implementation outcomes of the intervention (acceptability, appropriateness, feasibility and fidelity). We will also explore the detailed process and content perspectives of how participants received the intervention. Approximately 24 purposively selected participants from the intervention arm, balanced by gender (women/men), age (60-69/≥70), and PHQ-9 score at first follow-up (0-4, 5-9, ≥10), will be interviewed individually by telephone 1-4 weeks after the three-month follow-up. Trained research assistants, blinded to participant age and PHQ-9 score at inclusion and follow-up, will conduct these interviews using interview guides previously developed and tested by the research group. The interviews will cover the following topics related to participants and their perceptions of the programme: emotional state before and after the programme; experience of receiving and responding to messages; appropriateness, acceptability, engagement and fidelity of programme delivery; and the role of the participant’s support network. Interviews will last approximately 30 to 60 minutes.

Interviews will be transcribed verbatim. Transcriptions will be analysed in ATLAS.ti (version 23.0.0) using both deductive (using pre-established categories of acceptability, appropriateness, feasibility, and fidelity) and inductive (for other relevant categories that emerge from the analysis) approaches [51,52]. In-depth exploration of participants’ experiences will allow us to generate hypotheses about the relationship between clinical and implementation outcomes. Results will be reported according to the Consolidated Criteria for Reporting Qualitative Research (COREQ) [53].

### Dissemination policy

We expect to publish and present the results of this trial in relevant scientific journals and conferences. The results will also be presented to stakeholders once data collection has been completed. Access to anonymous participant data and statistical coding will be made available to the public upon request 24 months after publication of the effectiveness results. Each request should be accompanied by a research proposal with defined objectives and a statistical analysis plan, which will be evaluated by the joint principal investigators.

### Ethics

This study was authorised by the Secretaria da Saúde do Município de Guarulhos and approved by the Ethics Committee of the Hospital das Clínicas da Faculdade de Medicina da Universidade de Sao Paulo – HCFMUSP (CAPPesq, ref: 4.144.603, first approval 9 July 2020). This is the protocol version 6.0, 24 March 2023. The statistical analysis plan can be found at https://figshare.com/s/33a83a1ad01751dcf38c. The trial was registered in the Registro Brasileiro de Ensaios Clínicos (ReBEC, ensaiosclinicos.gov.br) on 21 October 2021 (submitted on 03 August 2021), RBR-6c7ghfd. Recruitment of participants started in September 2021.

Verbal informed consent will be obtained by research assistants at two points before the start of the screening assessment and during the invitation to participate in the trial. Informed consent will also be obtained from participants invited to the process evaluation interview. The ethics committee has approved the use of verbal consent. Participants will be informed prior to verbal consent that non-identifiable data will be used for publication. Verbal consent and subsequent interviews will be audio recorded if participants agree.

Anonymity will be guaranteed during data management and analysis. A member of the research team (GMO) will be responsible for extracting information on individuals from the list provided by the Guarulhos Health Secretariat and assigning random ID numbers to each one of them. The same number will identify the participants in the REDCap platform and the system developed to deliver the messages. The files containing the audio recordings of the interviews will be named with the corresponding ID number and the initials of the participants and stored in a secure online platform. Access to these systems (REDCap, message delivery system and audio file storage system) will be password protected and restricted according to the roles and permissions assigned to the research team members. Only one member of the research team (CAN) will have access to information that could identify individual participants during or after data collection. We will not keep paper records.

The risk of harm associated with the RCT and the intervention is considered to be minimal. We will not interfere with any pharmacological or non-pharmacological treatment that participants may be receiving during the trial. No relevant concomitant treatment will be prohibited during the trial. Acute suicide risk will be assessed using a standardised protocol whenever the ninth question of the PHQ-9 is scored 1 or higher at baseline or follow-up assessments. This protocol was successfully used in our previous RCT (PROACTIVE study) [24]. Once participants at acute suicide risk are identified, the research team will contact the UBS and, if possible, a family member.

### Oversight and monitoring

The Coordinating Centre and the composition of the Trial Steering Committee (TSC) are provided in S2 File. The TSC will discuss any relevant protocol modifications, and the Guarulhos health system managers and coordinators will be consulted as necessary. Changes to the study protocol will be submitted to the Ethics Committee (CAPPesq) for approval. The TSC will be informed about the progress of the trial. They will receive reports indicating whether the two arms are reasonably well balanced.

## Discussion

The prevention of depression is an important public health issue worldwide. One of the goals of prevention of major depression is to reduce depressive symptoms in older adults with subthreshold depression [54]. However, evidence of feasible, simple, low-cost psychosocial interventions for this population in primary care is lacking [9,55]. There is also a paucity of evidence on how best to deliver self-help interventions to this population. A recent review of eight trials that evaluated self-help interventions for subthreshold depression found that this strategy significantly reduced depressive symptoms [55]. Nevertheless, none of these trials used a fully automated messaging system, without the involvement of health professionals, to deliver the psychosocial intervention. We are aware of only two trials of fully automated digital interventions for subthreshold depression in adults. They found small to moderate effectiveness, but included also younger adults and different approaches and contents compared to PRODIGITAL, and were conducted in high-income countries [56,57]. The Viva Vida programme therefore aims to address some of these gaps in this area of research. Viva Vida’s automated delivery method via WhatsApp was chosen because it can facilitate the provision of care in settings where there is little or no availability of mental health services for vulnerable and isolated older people. It can also be useful at times when social distancing is required, such as during the COVID-19 pandemic, when people should limit social contact and stay at home. Additionally, WhatsApp is the most widely used messaging system in Brazil [58]. The remote delivery and the use of the storytelling technique allow older adults to participate regardless of literacy level and mobility ability, barriers to treatment that are common among older adults in LMICs. Finally, we plan to conduct economic and process evaluation analyses to provide evidence on the cost-effectiveness, acceptability, and feasibility of self-help digital psychosocial interventions for older adults with subthreshold depressive symptoms. If this evidence shows that the Viva Vida programme is low-cost, acceptable and effective for older adults with subthreshold depressive symptomatology in primary care settings, it could be implemented in the Brazilian Unified Health System.

## Supporting information

SPIRIT 2013 Checklist

Administrative information

PRODIGITAL protocol approved by the ethics committee - English

Viva Vida programme

## Data Availability

No datasets were generated or analysed during the current study. All relevant data from this study will be made available upon study completion.

## Acknowledgements

We would like to acknowledge the contribution of the staff of the UBSs in Guarulhos, Maria de Jesus Assis Ribeiro, Marcelo Bueno da Silva and other members of the Escola SUS-Guarulhos and the Guarulhos Health Secretariat who supported the development of the study. We would also like to thank Prof. David Ekers for his support in the development of the intervention.

This study was funded by the São Paulo Research Foundation (process number 2017/50094–2) and the Joint Global Health Trials initiative, jointly funded by the Department of Health and Social Care (DHSC), the Foreign, Commonwealth & Development Office (FCDO), the Medical Research Council (MRC) and Wellcome (process number MR/R006229/1). MS is supported by the CNPq-Brazil (307579/2019-0). FAPESP supported CAN (2018/19343-9 and 2022/05107-7), TVND (2021/04493-8), FAM (2020/02272-1), MOC (2020/14768-1), CHQS (2020/14504-4), GMO (2021/04230-7), MSS (2021/10148-1 and 2022/08668-0) and MMSM (2021/03849-3).

## Supporting information

**S1 File. SPIRIT checklist.** (DOCX)

**S2 File. Administrative information according to the SPIRIT checklist.** (DOCX)

**S3 File. Protocol that was approved by the ethics committee, in Brazilian Portuguese (original).** (DOCX)

**S4 File. Protocol that was approved by the ethics committee, in English (translation).** (DOCX)

**S5 File. Content of the messages of the Viva Vida programme.** (DOCX)

## References

[1] Meeks T, Vahia I, Lavretsky H, Kulkarni G, Jeste D. A tune in “a minor” can “b major”: A review of epidemiology, illness course, and public health implications of subthreshold depression in older adults. J Affect Disord. 2011;129: 126–142. doi:10.1016/j.jad.2010.09.015.

[2] Cherubini A, Nisticò G, Rozzini R, Liperoti R, Di Bari M, Zampi E, et al. Subthreshold depression in older subjects: an unmet therapeutic need. J Nutr Health Aging. 2012;16: 909–913. doi:10.1007/s12603-012-0373-9.

[3] Kroenke K. When and how to treat subthreshold depression. JAMA. 2017;317: 702–704. doi:10.1001/jama.2017.0130.

[4] Rodríguez MR, Nuevo R, Chatterji S, Ayuso-Mateos JL. Definitions and factors associated with subthreshold depressive conditions: a systematic review. BMC Psychiatry. 2012;12: 181. doi:10.1186/1471-244X-12-181.

[5] Kroenke K. Minor depression: midway between major depression and euthymia. Ann Intern Med. 2006;144: 528–530. doi:10.7326/0003-4819-144-7-200604040-00013.

[6] Ludvigsson M, Marcusson J, Wressle E, Milberg A. Morbidity and mortality in very old individuals with subsyndromal depression: an 8-year prospective study. Int Psychogeriatr. 2019;31: 1569–1579. doi:10.1017/S1041610219001480.

[7] Wiktorsson S, Runeson B, Skoog I, Östling S, Waern M. Attempted suicide in the elderly: characteristics of suicide attempters 70 years and older and a general population comparison group. Am J Geriatr Psychiatry. 2010;18: 57–67. doi:10.1097/JGP.0b013e3181bd1c13.

[8] Cuijpers P, Vogelzangs N, Twisk J, Kleiboer A, Li J, Penninx BW. Differential mortality rates in major and subthreshold depression: meta-analysis of studies that measured both. Br J Psychiatry. 2013;202: 22– 27. doi:10.1192/bjp.bp.112.112169.

[9] Lee SY, Franchetti MK, Imanbayev A, Gallo JJ, Spira AP, Lee HB. Non-pharmacological prevention of major depression among community-dwelling older adults: a systematic review of the efficacy of psychotherapy interventions. Arch Gerontol Geriatr. 2012;55: 522–529. doi:10.1016/j.archger.2012.03.003.

[10] van’t Veer-Tazelaar PJ, van Marwijk HWJ, van Oppen P, van Hout HPJ, van der Horst HE, Cuijpers P, et al. Stepped-care prevention of anxiety and depression in late life: a randomized controlled trial. Arch Gen Psychiatry. 2009;66: 297–304. doi:10.1001/archgenpsychiatry.2008.555.

[11] Spek V, Cuijpers P, Nyklíček I, Smits N, Riper H, Keyzer J, et al. One-year follow-up results of a randomized controlled clinical trial on internet-based cognitive behavioural therapy for subthreshold depression in people over 50 years. Psychol Med. 2008;38: 635–639. doi:10.1017/S0033291707002590.

[12] Ciechanowski P, Wagner E, Schmaling K, Schwartz S, Williams B, Diehr P, et al. Community-integrated home-based depression treatment in older adults: a randomized controlled trial. JAMA. 2004;291: 1569–1577. doi:10.1001/jama.291.13.1569.

[13] Williams JW, Barrett J, Oxman T, Frank E, Katon W, Sullivan M, et al. Treatment of dysthymia and minor depression in primary care: a randomized controlled trial in older adults. JAMA. 2000;284: 1519–1526. doi:10.1001/jama.284.12.1519.

[14] Gilbody S, Lewis H, Adamson J, Atherton K, Bailey D, Birtwistle J, et al. Effect of collaborative care vs usual care on depressive symptoms in older adults with subthreshold depression: The CASPER randomized clinical trial. JAMA. 2017;317: 728–737. doi:10.1001/jama.2017.0130.

[15] Riadi I, Kervin L, Dhillon S, Teo K, Churchill R, Card KG, et al. Digital interventions for depression and anxiety in older adults: a systematic review of randomised controlled trials. Lancet Healthy Longev. 2022;3: e558–e571. doi:10.1016/S2666-7568(22)00121-0.

[16] Witteveen AB, Young S, Cuijpers P, Ayuso-Mateos JL, Barbui C, Bertolini F, et al. Remote mental health care interventions during the COVID-19 pandemic: An umbrella review. Behaviour Research and Therapy. 2022;159. doi:10.1016/j.brat.2022.104226.

[17] Brasil, Ministério da Saúde. Renda média domiciliar per capita. In: DATASUS [Internet]. 2010 [cited 5 Nov 2023]. Available: https://datasus.saude.gov.br/trabalho-e-renda-censos-1991-2000-e-2010.

[18] Brasil, Instituto Brasileiro de Geografia e Estatística (IBGE). Guarulhos. In: Panorama Censo [Internet]. 2022 [cited 26 Nov 2023]. Available: https://censo2022.ibge.gov.br/panorama/.

[19] Brasil, Ministério da Saúde. Taxa de analfabetismo. In: DATASUS [Internet]. 2010 [cited 22 Mar 2023]. Available: https://datasus.saude.gov.br/educacao-censos-1991-2000-e-2010.

[20] Brasil, Instituto Brasileiro de Geografia e Estatística (IBGE). Percentual de pessoas que tinham telefone móvel celular para uso pessoal na população de 10 anos ou mais de idade, por sexo e grupo de idade. In: SIDRA [Internet]. 2021 [cited 5 Nov 2023]. Available: https://sidra.ibge.gov.br/tabela/7361.

21. Brasil, Instituto Brasileiro de Geografia e Estatística (IBGE). Percentual de pessoas que utilizaram a Internet no período de referência dos últimos três meses na população de 10 anos ou mais de idade, por grupo de idade. In: SIDRA [Internet]. 2021 [cited 5 Nov 2023]. Available: https://sidra.ibge.gov.br/tabela/7334.

[22] Kroenke K, Spitzer RL, Williams JBW. The PHQ-9: validity of a brief depression severity measure. J Gen Intern Med. 2001;16: 606–613. doi:10.1046/j.1525-1497.2001.016009606.x.

[23] Nakamura CA, Scazufca M, Peters TJ, Fajersztajn L, Van de Ven P, Hollingworth W, et al. Depressive and subthreshold depressive symptomatology among older adults in a socioeconomically deprived area in Brazil. Int J Geriatr Psychiatry. 2022;37. doi:10.1002/gps.5665.

[24] Scazufca M, Nakamura CA, Peters TJ, Henrique MG, Seabra A, La Rotta EG, et al. A collaborative care psychosocial intervention to improve late life depression in socioeconomically deprived areas of Guarulhos, Brazil: the PROACTIVE cluster randomised controlled trial protocol. Trials. 2020;21: 914. doi:10.1186/s13063-020-04826-w.

[25] Scazufca M, Nakamura CA, Seward N, Moreno-Agostino D, Van de Ven P, Hollingworth W, et al. A task-shared, collaborative care psychosocial intervention for improving depressive symptomatology among older adults in a socioeconomically deprived area of Brazil (PROACTIVE): a pragmatic, two-arm, parallel-group, cluster-randomised controlled trial. Lancet Healthy Longev. 2022;3: e690–e702. doi:10.1016/S2666-7568(22)00194-5.

[26] Schotte CKW, Van Den Bossche B, De Doncker D, Claes S, Cosyns P. A biopsychosocial model as a guide for psychoeducation and treatment of depression. Depress Anxiety. 2006;23: 312–324. doi:10.1002/da.20177.

[27] Kanter JW, Manos RC, Bowe WM, Baruch DE, Busch AM, Rusch LC. What is behavioral activation? A review of the empirical literature. Clin Psychol Rev. 2010;30: 608–620. doi:10.1016/j.cpr.2010.04.001

[28] Krause RJ, Rucker DD. Strategic storytelling: when narratives help versus hurt the persuasive power of facts. Pers Soc Psychol Bull. 2020;46: 216–227. doi:10.1177/0146167219853845.

[29] Carswell K, Harper-Shehadeh M, Watts S, Van’t Hof E, Ramia JA, Heim E, et al. Step-by-Step: a new WHO digital mental health intervention for depression. Mhealth. 2018;4: 34. doi:10.21037/mhealth.2018.08.01.

[30] Mehrotraa S, Tripathib R. Recent developments in the use of smartphone interventions for mental health. Curr Opin Psychiatry. 2018;31: 379–388. doi:10.1097/YCO.0000000000000439.

[31] Chen YRR, Schulz PJ. The effect of information communication technology interventions on reducing social isolation in the elderly: a systematic review. J Med Internet Res. 2016;18: e18. doi:10.2196/jmir.4596.

[32] National Institute for Health and Care Excellence. Depression in adults: treatment and management. 29 Jun 2022 [cited 22 Mar 2023]. Available: https://www.nice.org.uk/guidance/ng222/resources/depression-in-adults-treatment-and-management-pdf-66143832307909.

[33] Gelenberg AJ, Freeman MP, Markowitz JC, Rosenbaum JF, Thase ME, Trivedi MH, et al. Practice guideline for the treatment of patients with major depressive disorder. American Psychiatric Association; 2010. Available: https://psychiatryonline.org/pb/assets/raw/sitewide/practice_guidelines/guidelines/mdd-1410197717630.pdf.

[34] Spitzer RL, Kroenke K, Williams JB, Löwe B. A brief measure for assessing generalized anxiety disorder: the GAD-7. Arch Intern Med. 2006;166: 1092–1097. doi:10.1001/archinte.166.10.1092.

[35] Hughes ME, Waite LJ, Hawkley LC, Cacioppo JT. A short scale for measuring loneliness in large surveys: results from two population-based studies. Res Aging. 2004;26: 655–672. doi:10.1177/0164027504268574.

[36] Devlin NJ, Krabbe PF. The development of new research methods for the valuation of EQ-5D-5L. Eur J Health Econ. 2013;14: S1–S3. doi:10.1007/s10198-013-0502-3.

[37] Grewal I, Lewis J, Flynn T, Brown J, Bond J, Coast J. Developing attributes for a generic quality of life measure for older people: preferences or capabilities? Soc Sci Med. 2006;62: 1891–1901. doi:10.1016/j.socscimed.2005.08.023.

[38] Scazufca M, Couto MCP de P, Henrique MG, Mendes AV, Matijasevich A, Pereda PC, et al. Pilot study of a two-arm non-randomized controlled cluster trial of a psychosocial intervention to improve late life depression in socioeconomically deprived areas of São Paulo, Brazil (PROACTIVE): feasibility study of a psychosocial intervention for late life depression in São Paulo. BMC Public Health. 2019;19: 1152. doi:10.1186/s12889-019-7495-5.

[39] Nakamura CA, Scazufca M, Moretti FA, Didone TVN, Martins MM de S, Pereira LA, et al. Digital psychosocial intervention for depression among older adults in socioeconomically deprived areas in Brazil (PRODIGITAL-D): protocol for an individually randomised controlled trial. Trials. 2022;23: 761. doi:10.1186/s13063-022-06623-z.

[40] Harris PA, Taylor R, Thielke R, Payne J, Gonzalez N, Conde JG. Research electronic data capture (REDCap)--a metadata-driven methodology and workflow process for providing translational research informatics support. J Biomed Inform. 2009;42: 377–381. doi:10.1016/j.jbi.2008.08.010.

[41] Harris PA, Taylor R, Minor BL, Elliott V, Fernandez M, O’Neal L, et al. The REDCap consortium: building an international community of software platform partners. J Biomed Inform. 2019;95: 103208. doi:10.1016/j.jbi.2019.103208.

[42] Schulz KF, Altman DG, Moher D, CONSORT Group. CONSORT 2010 statement: updated guidelines for reporting parallel group randomised trials. PLoS Med. 2010;7: e1000251. doi:10.1371/journal.pmed.1000251.

[43] Peugh JL, Strotman D, McGrady M, Rausch J, Kashikar-Zuck S. Beyond intent to treat (ITT): A complier average causal effect (CACE) estimation primer. J Sch Psychol. 2017;60: 7–24. doi:10.1016/j.jsp.2015.12.006.

[44] Lee KJ, Carlin JB. Multiple imputation for missing data: fully conditional specification versus multivariate normal imputation. Am J Epidemiol. 2010;171: 624–632. doi:10.1093/aje/kwp425.

[45] Sterne JA, White IR, Carlin JB, Spratt M, Royston P, Kenward MG, et al. Multiple imputation for missing data in epidemiological and clinical research: potential and pitfalls. BMJ. 2009;338: b2393. doi:10.1136/bmj.b2393.

[46] Rubin D. Multiple Imputation for Nonresponse in Surveys. Wiley Series in Probability and Statistics. New York: John Wiley & Sons, Inc.; 1987.

[47] Carpenter JR, Kenward MG, White IR. Sensitivity analysis after multiple imputation under missing at random: A weighting approach. Stat Methods Med Res. 2007;16: 259–275. doi:10.1177/0962280206075303.

[48] Carpenter J, Pocock S, Lamm CJ. Coping with missing data in clinical trials: a model-based approach applied to asthma trials. Stat Med. 2002;21: 1043–1066. doi:10.1002/sim.1065.

[49] Héraud-Bousquet V, Larsen C, Carpenter J, Desenclos J-C, Le Strat Y. Practical considerations for sensitivity analysis after multiple imputation applied to epidemiological studies with incomplete data. BMC Med Res Methodol. 2012;12: 73. doi:10.1186/1471-2288-12-73.

[50] Williams A, Kind P. The present state of play about QALYs. In: Hopkins A, editor. Measures of the quality of life and the uses to which such measures may be put. London: RCP Publications; 1992.

[51] Braun V, Clarke V. Using thematic analysis in psychology. Qual Res Psychol. 2006;3: 77–101. doi:10.1191/1478088706qp063oa.

[52] Green J, Thorogood N. Qualitative Methods for Health Research. 4th ed. Sage; 2004.

[53] Tong A, Sainsbury P, Craig J. Consolidated criteria for reporting qualitative research (COREQ): a 32-item checklist for interviews and focus groups. Int J Qual Health Care. 2007;19: 349–357. doi:10.1093/intqhc/mzm042.

[54] Cuijpers P, Smit F, Van Straten A. Psychological treatments of subthreshold depression: a meta-analytic review. Acta Psychiatr Scand. 2007;115: 434–441. doi:10.1111/j.1600-0447.2007.00998.x.

[55] Corpas J, Gilbody S, McMillan D. Cognitive, behavioural or cognitive-behavioural self-help interventions for subclinical depression in older adults: a systematic review and meta-analysis. J Affect Disord. 2022;308: 384–390. doi:10.1016/j.jad.2022.04.085.

[56] Spek V, Nyklíček I, Smits N, Cuijpers P, Riper H, Keyzer J, et al. Internet-based cognitive behavioural therapy for subthreshold depression in people over 50 years old: A randomized controlled clinical trial. Psychol Med. 2007;37: 1797–1806. doi:10.1017/S0033291707000542.

[57] Morgan AJ, Jorm AF, Mackinnon AJ. Email-based promotion of self-help for subthreshold depression: Mood Memos randomised controlled trial. British Journal of Psychiatry. 2012;200: 412–418. doi:10.1192/bjp.bp.111.101394.

[58] Newman N, Fletcher R, Robertson CT, Eddy K, Kleis Nielsen R. Reuters Institute Digital News Report 2022. Reuters Institute for the Study of Journalism; 2022. Available: https://reutersinstitute.politics.ox.ac.uk/sites/default/files/2022-06/Digital_News-Report_2022.pdf.

