## Supplementary material for "Self-help digital psychosocial intervention for older adults with subthreshold depressive symptoms in primary care in Brazil (PRODIGITAL): Protocol for an individually randomised controlled trial": Administrative information

**Supplementary information file 2**

**Administrative information according to SPIRIT checklist (items 1 to 5)**

| Title | Self-help digital psychosocial intervention for older adults with subthreshold depressive symptoms in primary care in Brazil (PRODIGITAL): Protocol for an individually randomised controlled trial |
| --- | --- |
| Trial registration | The protocol is registered with Registro Brasileiro de Ensaios Clinicos (ReBEC), number RBR-6c7ghfd. |
| Protocol version | Version 6.0, 24 March 2023. |
| Funding | This study was funded by the São Paulo Research Foundation (process number 2017/50094–2) and the Joint Global Health Trials initiative, jointly funded by the Department of Health and Social Care (DHSC), the Foreign, Commonwealth & Development Office (FCDO), the Medical Research Council (MRC) and Wellcome (process number MR/R006229/1). MS is supported by the CNPq-Brazil (307579/2019-0). FAPESP supported CAN (2018/19343-9 and 2022/05107-7), TVND (2021/04493-8), FAM (2020/02272-1), MOC (2020/14768-1), CHQS (2020/14504-4), GMO (2021/04230-7), MSS (2021/10148-1 and 2022/08668-0) and MMSM (2021/03849-3). The funders did not and will not have a role in study design, data collection and analysis, the decision to publish, or the preparation of the manuscript. There was no additional external funding received for this study. |
| Names, affiliations, and roles of protocol contributors | Authors: Thiago Vinicius Nadaleto Didone^1,2^, Carina Akemi Nakamura^1^, Nadine Seward^3^, Felipe Azevedo Moretti^1^, Monica Souza dos Santos^1^, Mariana Mendes de Sá Martins^1^, Luara Aragoni Pereira^4^, Evelyn da Silva Bitencourt^5^, Marcelo Oliveira da Costa^1^, Caio Hudson Queiroz de Souza^1^, Gabriel Macias de Oliveira^1^, Marcelo Machado^6^, Jamie Murdoch^7^, Pepijn Van de Ven^8^, William Hollingworth^9^, Tim J. Peters^10^, Ricardo Araya^3^, Marcia Scazufca^1,4^  Affiliations: **^1^**Departamento de Psiquiatria, Faculdade de Medicina FMUSP, Universidade de Sao Paulo, Sao Paulo, SP, Brazil; ^2^Departamento de Saúde Coletiva, Centro de Ciências da Saúde, Universidade Estadual de Londrina, Londrina, PR, Brazil; ^3^ Health Service and Population Research, Institute of Psychiatry, Psychology and Neuroscience, King’s College London, London, United Kingdom; ^4^Instituto de Psiquiatria, Hospital das Clinicas HCFMUSP, Faculdade de Medicina, Universidade de Sao Paulo, Sao Paulo, SP, Brazil; ^5^Faculdade de Arquitetura e Urbanismo FAU, Universidade de Sao Paulo, Sao Paulo, SP, Brazil; ^6^Debasé Audiovisual, Sao Paulo, SP, Brazil; ^7^Department of Population Health Sciences, King’s College London, London, United Kingdom; ^8^Health Research Institute, University of Limerick, Limerick, Ireland; ^9^Health Economics Bristol, Population Health Sciences, Bristol Medical School, University of Bristol, Bristol, United Kingdom; ^10^Population Health Sciences, Bristol Medical School, and Bristol Dental School, University of Bristol, Bristol, United Kingdom  Roles: Marcia Scazufca and Ricardo Araya are the principal investigators; they led the proposal, conceived the study design, and developed the study protocol and process evaluation plan. Tim J Peters and William Hollingworth are co-investigators; they conceived the study design and developed the study protocol. Thiago VN Didone is the intervention coordinator; he developed the study protocol. Carina A Nakamura is the research coordinator; she conceived the study design and developed the study protocol. Felipe A Moretti coordinated and conceived the information technology plan. Nadine Seward conceived the analysis plan. Caio HQ de Souza, Gabriel M de Oliveira, Marcelo O da Costa and Pepijn Van de Ven developed the information technology plan. Marcelo Machado, Luara A Pereira e Mariana M de S Martins support the development of the intervention. Monica S dos Santos, Evelyn da S Bitencourt and Jamie Murdoch designed the process evaluation plan. Thiago VN Didone, Carina A Nakamura, Nadine Seward and Marcia Scazufca drafted the manuscript. |
| Name and contact information  for the trial sponsor | Sponsors: Universidade de Sao Paulo (USP), Brazil, and King’s College London (KCL), United Kingdom  Principal Investigators contact: Dr Marcia Scazufca, Faculdade de Medicina da Universidade de Sao Paulo, Brazil,; Professor Ricardo Araya, King’s College London, United Kingdom, |
| Role of study sponsor and funders | The sponsors and funders did not and will not have a role in study design, data collection and analysis, the decision to publish, or the preparation of the manuscript. |
| Composition, roles, and responsibilities of the coordinating centre, steering committee, endpoint adjudication committee, data management team, and other individuals or groups overseeing the trial, if applicable | Coordinating centre: Universidade de Sao Paulo, Sao Paulo, Brazil  Trial Steering Committee (TSC):   1) Independent members: Professor Simon Gilbody (University   of York, United Kingdom); Professor Roberto Alves Lourenço (Universidade Católica do Rio de Janeiro, Brazil); Dr Rodrigo Fonseca Martins Leite (Universidade Municipal de São Caetano do Sul, Brazil); and Paula Verônica Martini Maciel (Escola SUS, Guarulhos Secretary of Health, Brazil).   2) Trial team members: Dr Marcia Scazufca (Universidade   de Sao Paulo, Brazil); Professor Ricardo Araya (King’s College London, United Kingdom); and Professor Tim Peters (University of Bristol, United Kingdom).  The Trial Steering Committee (TSC) will meet once a year, either virtually or in person. |
