## Supplementary material for "Self-help digital psychosocial intervention for older adults with subthreshold depressive symptoms in primary care in Brazil (PRODIGITAL): Protocol for an individually randomised controlled trial": PRODIGITAL protocol approved by the ethics committee - English

**Supplementary information file 4**

**PRODIGITAL protocol approved by the ethics committee in English (translation from the original version)**

**Digital health communication strategies for reducing depressive symptoms in older adults: randomised controlled trial nested in the "Care programme for older people with depressive symptoms in primary care", FAPESP Case 2017/50094-2**

Digital care programme for older people with mild symptoms of depression

Responsible Researcher: Marcia Scazufca

Institute of Psychiatry, University of São Paulo Medical School

2020

**Abstract**

We are proposing to conduct a randomised controlled clinical trial with older adults with subclinical symptoms of depression to test the effectiveness and cost-effectiveness of a digital intervention (intervention group) for the reduction of depressive symptoms compared to the control group (two messages) (PRODIGITAL). PRODIGITAL is a subproject of the study "Care programme for older adults with depressive symptoms in primary care: cluster randomised controlled trial for depression in older adults residents in socioeconomically disadvantaged areas of Sao Paulo, Brazil (PROACTIVE) ". We estimate that with the inclusion of 450 participants (225 in each group) it will be possible to find a difference in the mean score of the assessment of depressive symptoms with the Patient Health Questionnaire-9 (PHQ-9) (primary outcome) of at least 0.33 standard deviations lower in the intervention group than in the control group, three months after inclusion in PRODIGITAL (statistical power of 80-85% and alpha of 0.05%). The main secondary endpoints are to assess the cost-effectiveness of the digital intervention compared to the control group and to examine whether the gains regarding decreased depressive symptom intensity observed at three months are maintained at five months after study inclusion. The participants of the present clinical trial will be recruited simultaneously to the study "Digital psychosocial intervention for older adults with depression living in low-income areas of Guarulhos, São Paulo: Randomised clinical trial (PRODIGTAL-D)", which will include older adults with depression. The older adults aged 60 years or older, registered in the 24 primary care clinics of the municipality of Guarulhos, and who in the screening evaluation present subclinical symptoms of depression (5≤PHQ-9≤9), and have a mobile phone to receive messages from the WhatsApp application, will be eligible for PRODIGITAL. The individual randomization of the participants will be done remotely and after the end of the inclusion procedures. After randomization, the participants allocated to the control group will receive routine care of the primary care clinics and two messages through the WhatsApp application on their mobile phones, with information on the main signs of depression and suggestions of some activities that can help them improve mood. The intervention group will receive the routine care and the digital psychosocial intervention, through sending audio messages or 'memes' four times a week, by WhatsApp application, for a period of six weeks. The messages will have motivational content based on psychoeducation and behavioural activation techniques that aim to promote positive changes in lifestyle and social interactions of the participants. Before the start of the trial, we will conduct a pilot study with ten aged people to assess possible difficulties in understanding and receiving the messages. All the contacts of the researchers with the study participants will be made by telephone. Analyses will be performed according to the CONSORT guidelines. Recruitment (screening and baseline assessments) and inclusion of participants will be carried out after participants have been informed about the objectives of the study, read the Informed Consent Form and consent to participate. This project does not present significant risks for the participants.

**Acronyms used in the project**

CEP/FMUSP – Research Ethics Committee of the University of São Paulo Medical School

CAPPesq – Ethics Committee for the Analysis of Research Projects of the Hospital das Clínicas of the University of São Paulo Medical School

EqSF – Family Health Team

ESF – Family Health Strategy

PHQ-2 – Patient Health Questionnaire-2

PHQ-9 – Patient Health Questionnaire-9

PROACTIVE – Randomised controlled trial for the treatment of older people with depression

PRODIGITAL – Randomised controlled trial in older people with subclinical symptoms of depression for the reduction of depressive symptoms

PRODIGITAL-D – Digital psychosocial intervention for older adults with depression living in low-income areas of Guarulhos, São Paulo: randomised clinical trial

PRSI – Immediate Suicide Risk Protocol

QALYs – Quality Adjusted Life Years

SUS – Unified Health System

TCLE – Free and Informed Consent Form

UBS – Primary Care Clinic

WhatsApp – A multi-platform [instant mess](https://pt.wikipedia.org/wiki/Mensageiro_instant%C3%A2neo)aging and calling app for text, voice, images, videos and PDF documents for [smartphones](https://pt.wikipedia.org/wiki/Smartphone), as well as making free calls over an internet connection.

**Introduction**

The presence of mild depressive symptoms and of insufficient severity for the diagnosis of depression (subclinical depression) is an important risk factor for the incidence of depression and deterioration of quality of life in older adults.^1-3^ Studies with populations in different countries have found incidence rates of depression in older adults ranging from 7.5% to 30.6%.^4-6^ Despite this being a significant problem for the health of older adults, there are no clear guidelines, based on scientific evidence, of simple and effective interventions that can be used with primary health care services users.

A Cochrane systematic review showed that digital health communication strategies can be great tools to promote greater autonomy and proactivity in individuals, improvement in social interaction, reduction of hopelessness, greater knowledge about the management of specific conditions, expansion of behavioural strategies and better clinical outcomes.^7-8^ However, most research on digital solutions conducted with individuals with specific clinical outcomes are still categorized as of low or moderate quality, suggesting the need to conduct new studies.^8^

As part of the studies that make up the "Care programme for older people with depressive symptoms in primary care - (PROACTIVE) ", approved by the Research Ethics Committee of the University of São Paulo Medical School (CEP/FMUSP) on 22 August 2018 (CAAE: 86168318.3.0000.0065, opinion no. 2.836.569), we are proposing to develop and test in a randomised controlled clinical trial an innovative digital intervention that aims to reduce the intensity of depressive symptoms in older adults with subclinical symptoms of depression living in socioeconomically deprived areas of the municipality of Guarulhos (PRODIGITAL). FAPESP granted on 21 November 2019 an addendum to the PROACTIVE funding (FAPESP Case 2017/50094-2) to carry out PRODIGITAL. The Health Secretariat of the municipality of Guarulhos approved PRODIGITAL on 09 January 2020.

**Hypothesis**

The difference in the mean score of the Patient Health Questionnaire-9 (PHQ-9),^9-12^ used for the assessment of depressive symptoms, will be at least 0.33 standard deviations lower in the intervention group than in the control group at three months after study inclusion, with 80-85% statistical power and significance level (alpha) of 0.05 in a two-tailed model. Such a difference is considered clinically significant for the reduction of depressive symptoms in the population.

**Objectives**

Primary

- To test the effectiveness of a digital intervention, by sending WhatsApp messages, to reduce the intensity of depression symptoms in older adults with subclinical depressive symptoms assisted by Family Health Teams (EqSF) in Primary Care Clinics (UBSs) in the municipality of Guarulhos, compared with guidance on depression by sending two mobile phone messages, three months after inclusion in the study.

Secondary

- To evaluate the cost-effectiveness of the digital intervention compared to guidance on depression by sending two mobile phone messages (control group);
- To examine whether the gains in decreasing the intensity of depressive symptoms observed at three months are maintained at five months after inclusion in the study;
- To investigate by means of qualitative methods the possible mechanisms of operation of the digital intervention.

**Method**

PRODIGITAL is a two-group, randomised controlled clinical trial with individual randomisation. This study is part of the "Care programme for older people with depressive symptoms in primary care". The remote psychosocial intervention of PRODIGITAL is based on the psychosocial intervention conducted in the PROACTIVE study. Before the start of PRODIGITAL, a pilot study will be conducted to test sending the messages for a group of older people with similar characteristics to the participants in the future clinical trial.

Pilot Study

The pilot study will be conducted before the start of the RCT and aims to test the messaging that will be used in the psychosocial intervention (PRODIGITAL). Ten individuals aged 60 years or older who use UBSs will be included. We will ask the managers of the UBSs of Guarulhos to help us in the selection of participants for the pilot study. We will use 12 of the 48 messages developed for the RCT. The messages will be sent for a period of three weeks (4 messages per week, voice messages and images), following the same protocol that will be used in the RCT. At the end of the three weeks, we will contact the participants of the pilot study by telephone to find out if they had any technical difficulties in receiving the messages, or difficulty in reading and understanding them. After being informed about the objectives and method of conducting the pilot study, informed consent will be sought from all participants (consent will be audio recorded).

Setting and participants

The PRODIGITAL study will include aged individuals registered with the same 24 UBSs of the municipality of Guarulhos participating in the PROACTIVE study.

Eligibility criteria for UBSs and Family Health Teams (EqSF) in PROACTIVE

- A list was obtained from the Health Secretariat (SMS) of Guarulhos with all the UBSs with EqSF in the municipality. The eligibility criteria of the UBSs were: UBSs with an average of at least 400 registered individuals older than 60 years per EqSF and at least four teams responsible for at least 300 individuals; authorization of the UBS manager to participate in the study;
- The study proposal was presented at a meeting attended by the managers (or their representatives) of the eligible UBSs. At this meeting it was explained that the participation of the UBS in the study was voluntary. All managers of eligible UBSs that agreed to participate sent a formal authorisation letter to the study coordinators. After the authorisation of the UBSs managers, 24 UBSs were randomly selected to participate in the study;
- Random selection of UBSs and EqSFs from each participating UBSs in PROACTIVE was conducted remotely by individuals not involved in the recruitment of UBSs.

Inclusion criteria for PRODIGITAL participants

- Individuals aged 60 years or older registered with the EqSF of the 24 UBSs in the municipality of Guarulhos participating in PROACTIVE;
- Older adults who, in the screening interview, present subclinical depression (PHQ-9 score^9-12^ between 5 and 9, and at least score 1 in the sum of questions 1 and 2 of this questionnaire);
- Older adults have a mobile device that can receive messages on the WhatsApp application.

Exclusion criteria for PRODIGITAL participants

The exclusion criteria for PRODIGITAL participants are the same as those used in the PROACTIVE study:

1. Terminal illness, total deafness, severe impaired vision or blindness, severe mental illness and inability to communicate or participate in assessment interviews (non-native speaker, cognitive impairment). This information will be collected from the older adults themselves and/or from people close to them and/or through interviewer observation during recruitment assessments (screening and baseline interview). In doubtful situations, the case will be discussed with the coordination of the study.

2. Risk of suicide: Individuals who present at risk of suicide according to the 'Immediate Suicide Risk Protocol' (PRSI) will not be included in the study.

Assessments

The assessments described below will be conducted at recruitment and follow-up assessments (three and five months after study inclusion) and are the same as in the PRODIGITAL-D study. All assessments will be conducted by trained, independent research assistants (interviewers who were not involved in the design of the project and who will not participate in the preparation and delivery of the remote psychosocial intervention) via telephone calls. Secondary health data will also be collected from the Guarulhos electronic health information systems.

1. Recruitment: This will comprise the participant's mobile phone 'inclusion' and 'validation' assessments. The inclusion assessment will have two parts, screening and baseline (or initial) assessment, which will be carried out consecutively, during the same interview and after the participant's consent. Older adults who meet the inclusion criteria (and do not meet the exclusion criteria) will answer the baseline assessment questions and will be invited to participate in the 'validation' interview, which will verify the older adult mobile phone contact. Validation can be done during the same interview or in a new interview if necessary. Exclusion criteria will be checked during the inclusion and validation assessments.

1.a. Inclusion - Screening: older adults who have a mobile phone with the possibility of internet connection to receive WhatsApp messages and score greater than or equal to 1 in the evaluation with the Patient Health Questionnaire 2 (PHQ-2)^9-13^ will have the depression assessment completed with the PHQ-9, using the 7 questions of this questionnaire that are not part of the PHQ-2. PHQ-2 is a questionnaire used to screen for depression and is composed of the first two questions of the PHQ-9.^9-13^

1.b Inclusion - Baseline assessment: it will be conducted with older people with a score of 5≤PHQ-9≤9 (subclinical depression and no risk of suicide in the two weeks prior to the baseline assessment). The following assessments will be conducted: 'General Quality of Life' will be assessed with the EQ-5D-5L (EuroQol),^14-18^ broader well-being with the 'Older adults Capability' instrument (ICECAP-O),^19^ anxiety with the GAD-7,^20^ loneliness with the 3-item UCLA questionnaire.^21^ A standardized questionnaire will be used to assess the demographic and economic profile (age, gender, level of education, income, marital status, work situation), self-reported physical health problems (diabetes, hypertension, cancer, disability, others), use of psychotropic medication, tobacco use and digital literacy.

1.c. 'Validation': a research assistant will contact the participant by telephone to make the validation of the mobile phone number, which includes sending a message to the participant's mobile phone using the WhatsApp application. In this interview the participant will be informed about all the procedures of the clinical trial and will be asked for Informed Consent.

**2.** Follow-up assessments: three and five months after inclusion: The PHQ-9 (assessment of the main outcome) and the other assessments carried out at recruitment (EuroQol, ICECAP-O, GAD-7, UCLA) will be repeated and possible changes in demographic and economic profile, physical health problems, use of psychotropic medication and use of alcohol and tobacco will be repeated. Use of health services and expenditure on medication in the period between inclusion and each of the follow-up assessments will also be assessed.

**3.** Process evaluation: The acceptability of the programme (intervention group) by the older adults will be done through descriptive analysis of engagement throughout the psychosocial intervention, measured by the percentage (%) of participants who received and viewed the contents of the messages. During the intervention, messages will be sent to assess the degree of satisfaction with the programme, using Likert scales (with variations from very satisfied to dissatisfied, for example), in order to capture the feelings and understanding of the communicational steps. These messages will also ask participants to send voice messages to the study sharing their experiences with the programme. Satisfaction scales will be sent at 6 separate times (once a week). Such a Likert scale model is often used in satisfaction analyses of programmes, services, or other mental health programmes.

Routine data existing in the electronic information systems of the Health System of Guarulhos (or in medical records if the information is not available in the electronic information systems) will be extracted to complement the information obtained with the participants: use of psychotropic medications (medication, dosage and schedule) and consultations with physicians and nurses in the UBSs. These data will be extracted regarding the period between the inclusion of the participant in the study (intervention and control group) to the date of the second follow-up assessment and will be stored in a safe environment (study server), under the responsibility of the researcher responsible for conducting the clinical trial. At the follow-up assessments, participants will be asked whether they had any hospital admissions (one night or more), referrals for specialized mental health care (CAPS or professionals), and whether they needed to pay for mental health-related care (medications, professionals etc.).

Sample Size

We estimate that it will take the inclusion of 142-162 participants in each group to obtain a difference of 0.33 SD between the two groups (mean PHQ-9 between 5.9 and 6.4 with standard deviation of 1.5 points based on data from previous studies by our research group) with 80-85% statistical power and alpha of 0.05 in a two-tailed model. This number needs to be inflated by 25% to account for possible loss to follow-up. For this reason, we will recruit a total of 225 participants in each group. Based on the PROACTIVE study, we believe that it will be feasible to include 450 participants in the 24 UBSs participating in the study. Data from the first phase of inclusion in the PROACTIVE study indicated that approximately 55% of older adults have a mobile device and use the WhatsApp app.

Inclusion and allocation of study participants

The allocation of participants to the control and intervention groups will be done by individual randomization, soon after each inclusion of the participants in the study. In the present study, cluster allocation will not be necessary as it is unlikely that the digital intervention will lead to contamination between groups. Before starting recruitment assessments, participants will be informed about the study and invited to participate. Recruitment of PRODIGITAL participants will be conducted simultaneously with that of PRODIGITAL-D participants. For this reason, the same Informed Consent Form (TCLE) will be used in both studies.

Planned interventions

The group of researchers participating in the conduct of the PROACTIVE study has already started the development of the remote intervention. Researchers specialised in developing digital technology for health interventions and professionals specialised in developing multimedia material are collaborating with this phase of the study. The additive granted by FAPESP for this study allowed the hiring of these professionals.

During the final phase of the intervention development, we will consult with older adults to obtain information about their acceptance of the intervention we are developing. We will ask managers of the UBSs participating in the study to nominate approximately 20 older adults registered in these units to participate in informal individual or small group discussions with the researchers responsible for developing the intervention. The observations made by the older adults during these conversations will be discussed by the researchers and may be used to improve aspects of the digital intervention pointed out as problematic by the older adults. The older adults will be informed about the research before we ask for their verbal informed consent, which will be audio recorded.

Remote psychosocial intervention: it will be based on concepts of interactive health communication applications. Participants will receive this intervention on their mobile phones through messages sent by the WhatsApp application. The way of carrying out the intervention (WhatsApp messages) was idealized to facilitate the participation of the older adults in the intervention, because it is of low cost and does not overburden health professionals. The content of these messages is being developed from the adaptation of psychoeducation and behavioural activation techniques, similar to those used in the PROACTIVE study.^22^ These contents will be presented to the participants using the storytelling technique. This technique has the ability to involve, convince, remind and motivate and is an important communication tool when you want to share and create bonds with other individuals and the potential to collaborate in adherence to the proposed intervention. We chose the use of this technique for its ability to improve the attention and retention of important information by the target audience, creating empathy and interest in a story, we can stimulate behaviours and the thoughts about problems that appear in other people.

Our proposal is to create two characters who go through similar situations to those of the target audience, which will facilitate the identification of the older adults with the characters. These characters will be played by an actor and an actress who will tell, through audio and text messages with images, their experiences with the day-to-day problems that we want to address in the intervention. The format of the messages will make this intervention accessible to older people who cannot read. These messages will show these characters' thoughts about their problems and how they deal with them, and will present our target audience with activities that have worked for the characters. The characters will give warnings about important health issues and give news to the audience about what happens in their daily routines and also changes that occur in the quality of their lives. Characters will use language appropriate to the chosen means of communication and vocabulary, tone of voice and manner of speaking appropriate for the target audience. The characters should be realistic enough to create empathy for the study participant when listening to or reading the messages, to enable the characters to get closer to the group. The stories of our characters will last six weeks and will be told by sending two messages a day (morning and afternoon) at a frequency of four times a week. This periodicity is considered ideal to maintain interest without tiring or becoming uninteresting. In this way, we believe it is possible to involve the target public and motivate them to pay attention and put into practice the suggested activities and techniques to improve mood related problems and consequently prevent depression. Another advantage of this proposal is that the participants will be able to look at the messages at any time of day, not depending on the opening hours of the UBSs. We will create a helpline for the participants. This helpline will only help participants who have technical difficulties using WhatsApp or who have difficulties receiving messages (change of phone number, disconnection of the line etc.). All participants' contacts with technical support will be noted (date and problem reported).

Control and intervention group: Soon after their inclusion in the study, the participants of the control group will receive a voice message and an image ('meme') with information about subclinical depressive symptoms, the importance of preventing these symptoms from worsening and where to seek care in the municipality of Guarulhos, should they need it. The participant will be advised to seek specialized mental health services or the UBSs in which he/she is registered.

Methodology of data analysis

The analyses and their presentation will be done according to the CONSORT guidelines.^23^ The primary analysis of comparison between the difference in means of the PHQ-9 of the two groups will be conducted by 'intention-to-treat' analysis. Analyses will first be conducted to verify that the groups are balanced with respect to their clinical and demographic characteristics. Multivariate mixed-effects regression models will be used to obtain the difference in the primary outcome between the two groups at three months, after adjusting analyses for PHQ-9 score at study inclusion and other study design characteristics. There will only be one primary analysis that will be conducted after the end of data collection. Secondary analyses will include comparison of the primary outcome assessment at three months (including the use of repeated measures regression models) and it will be repeated the analysis for the primary outcome adjusting for any observed imbalance between groups of potential predictor variables at the level of the individual and/or UBS (introduction of the latter would imply the use of appropriate multilevel models). We will also compare individuals according to the intervention they actually received (number of messages received and opened), accounting for any selection effects after random allocation. We will conduct exploratory subgroup analyses to investigate potential differential effects of the intervention according to pre-specified participant characteristics. These analyses will be interpreted cautiously, partly because we do not have a specific hypothesis for these results and partly because statistical power will be limited for interaction effects within the relevant regression models.^24^ We will also investigate the factors that may predict response to the intervention, using appropriate interaction terms in the regression models. The a priori hypotheses we seek to investigate are differential effects according to the individual's gender, age (continuous), years of education (continuous), presence of comorbidities, and severity of depressive symptoms (continuous PHQ-9 scores) at study inclusion.

Two approaches will be used for the economic evaluation: 1) The incremental cost-effectiveness ratio between the two study groups will be estimated using the primary clinical outcome measure (cost per patient recovered) and Quality Adjusted Life Years (QALYs)^14^ calculated using the EQ-5D-5L.^15^ These results will be presented as incremental cost-effectiveness ratios and cost-effectiveness acceptability curves, derived using bootstrapping techniques, which will show the likelihood of the intervention being cost-effective over a range of 'willingness-to-pay' levels. 2) The net monetary benefit statistic, using the difference in costs and the difference in QALYs between the two groups, will be calculated for different values of societal willingness-to-pay for a quality-of-life adjusted year. Sensitivity analysis will then be conducted to account for uncertainty in the assessments.

There will only be one main analysis that will be conducted after the completion of the clinical trial data collection. However, we will analyse the data collected at inclusion of study participants to provide important information to funding bodies on the progress of the study and as part of our strategy to start sensitising health managers for the future dissemination of the programme in the Brazilian Unified Health System.

Information collected in the recruitment phase (screening and baseline assessment) will be used to describe the profile of participants and estimate the prevalence of subclinical symptoms of depression and anxiety.

Information Technology (IT) Support System

Data collection and storage will be performed with the REDCap platform, hosted on the server of the USP Medical School. REDCap allows the study coordinators to follow the inclusion of the participants in the study practically in real time and to obtain information about the follow-up evaluations. Sending the collected data to the study server facilitates the evaluation of the quality of the information close to the collection, which allows the correction of any problems to be carried out.

Ethical Issues

The Health Secretariat of Guarulhos approved the present project (PRODIGITAL) on 09 January 2020. PRODIGITAL is a subproject of PROACTIVE and for this reason was sent to the CEP/FMUSP to be evaluated as an amendment to PROACTIVE. On the recommendation of Prof. Dr. Koike Folgueira, we are sending the project for evaluation by the Ethics Committee for the Analysis of Research Projects of the Hospital das Clinicas, University of São Paulo Medical School (CAPPesq) as a new project, for the following reasons: "The proposed changes were discussed in plenary session on 22 January 2020 and it was concluded that by the adoption of new intervention strategies in another study population, this subproject should not be submitted as an amendment but as a new project".

All evaluations will be carried out after obtaining the TCLE. The TCLE 1 will be used in the recruitment (screening and baseline assessment) of the participants. The TCLE 2 will be used in the second part of recruitment (validation), when the older adults will be invited to participate in the clinical trial. TCLE 3 will be used in the Pilot Study.

Risks

The clinical trial we are proposing does not present considerable risks to its participants. Research assistants will be trained for this study to maintain the confidentiality of the interviews and to deal with any discomfort participants may have during the assessments. The assessments that will be carried out are the same as those used in the PROACTIVE study (we have only excluded some questionnaires). No aged individual included in the PROACTIVE study reported any distress related to the assessments. The same exclusion criteria that were applied in the PROACTIVE study will be applied to the present study, as described above. These criteria exclude individuals with serious illness and/or disability from the study. Research assistants may discuss the contents of the interviews, when necessary, only with the study coordinators. In addition to explaining the purpose of the study and interviews before requesting consent to participate in the recruitment assessments, research assistants will reiterate to the older adults that the interviews are confidential and that participation in the study is not mandatory. Prior to the beginning of the recruitment, participants will be asked for consent for audio recording this interview. These recordings will be used to register the participant's consent and to do quality control of the interviews, specifically to assess whether the research assistant followed the protocol for informing the participant about the study to then seek consent. Our research group is experienced in conducting telephone interviews (PROACTIVE), none of the interviews conducted by telephone resulted in a health risk to the participant.

Participation in the study will not interfere in any way in the care of the EqSFs with the older adults or prevent general practitioners or other professionals in the team or other health services from initiating or changing any conduct for the health problems of the older adults included in the study.

Given the nature of the intervention, it is unlikely that participants will feel uncomfortable with the content of the audio or image/text messages they will receive on their mobile phones. The content of these messages will be based on psychoeducation and behavioural activation techniques, which are not considered harmful.^22^

Not often, but there is a possibility that some study participants report suicide risk in the study assessments, as depressive symptoms are a known risk factor for suicide. The PRSI was successfully applied in the PROACTIVE study (inclusion and follow-up), in assessments conducted in the participant's home and by telephone. In these interviews, every time the older adult scores at least 1 ('Several days') on the PHQ-9 question on suicidal ideation (9^th^ question) the suicide risk assessment is conducted. Suicide risk assessments will continue to be done in the same way and with the same protocol used in PROACTIVE. The PRSI is a 'Suicide Risk' identification protocol that contains instructions on what to do in cases where the risk is immediate. Older adults who show a risk of suicide in the screening assessment will not be invited to participate in the study and several actions will be taken during the interview to protect them. Research assistants are trained to carry out the assessment with the PRSI by telephone and to apply the necessary procedures if the aged individuals present an immediate risk of suicide. The main actions that should be taken in these situations are: 1. Explain to the older adult what is happening; 2. Do not end the interview until a relative or close person assumes the responsibility of caring for the older adult; 3. Explain to the relative or close person what is happening and how to proceed; 4. When the research assistant identifies an older adult at risk of suicide, the problem should be immediately informed to the coordinator of the study so that he contacts the UBS and informs the UBS manager of the situation. In the few situations that the application of the PRSI detected immediate suicide risk in PROACTIVE assessments (in assessments at the participant's home or by telephone), the recommendations of this protocol were followed by the researchers and the actions taken served to protect the older adults.

Benefits

The immediate benefit for the participants of PRODIGITAL will be the assessment of depressive symptoms, the possibility of receiving a minimal digital intervention, currently almost non-existent in health services, and guidance to seek health services if they perceive that their symptoms of depression are worsening. In the rare situations that interviewers detect that the older adult presents an immediate risk of suicide, we can consider that the actions that will be taken by the research assistants constitute a benefit. These situations are rarely identified by family members and health professionals and the actions of the research assistant are intended to decrease the risk that a suicide attempt will materialize and cause damage to health or death.

If the result of PRODIGITAL is positive, that is, the digital intervention proves to be cost-effective, the research group will work so that this simple, low-cost intervention that does not overburden health services will be incorporated by the Family Health Strategy (ESF) throughout the country and by other health services and thus benefit millions of older people who present subclinical depressive symptoms and do not receive support from the Unified Health System for these symptoms.

Other relevant information

Dr. Marcia Scazufca, the principal investigator of the PROACTIVE study, is also responsible for all stages of this study, which includes data collection and storage, supervision of the team, implementation of the intervention, analysis and dissemination of results and funding from FAPESP. The data collection and intervention of this study will be conducted exclusively in the 24 UBSs of the municipality of Guarulhos, where the clinical trial for depression treatment (PROACTIVE) was conducted.

The researchers collaborating on this study will support this project in their areas of academic expertise, i.e., the methodological and practical aspects of data collection and intervention implementation, data analysis and preparation of reports and scientific papers.

The recruitment of the older adults for the pilot study was performed with the same methodology that will be used for inclusion of participants in the clinical trial, i.e., telephone contact (mobile and landline) with the older adults registered in the UBSs of Guarulhos. One of the difficulties in conducting the pilot was to contact the older adults because many phones were inactive or did not answer our calls. After talking to the older adults who participated in the pilot study and UBS managers in Guarulhos, who are also contacting the older adults to call them to come and take the second dose of the COVID vaccine, we learned that the older adults usually do not answer their mobile phones when the number is unknown. One of the reasons for not answering the phone is the scams applied through phone calls that have been reported frequently in the press (TV, radio and newspapers). The recruitment stage is necessary to conduct the clinical trial. If this intervention is effective, the access to the Intervention Programme will not be in the same way as the recruitment for the clinical trial (telephone contact of researchers). Thus, the difficulty we are having recruiting participants for the clinical trial should not occur for the implementation of this programme in the 'real world'.

To improve the confidence of the older adults (and family members) registered with the UBSs (or other health services participating in the study) in relation to the study, we will implement the following actions (or similar actions in other health services participating in the study):

a. Put posters in the 24 participating UBSs with information about the study (objective of the study, sponsors, telephone number to contact those responsible for the study). Thus, the older adults who attend the UBS will be informed about the existence of the study and may contact us through a telephone number that will be dedicated specially to talk about possible doubts;

b. We will request that community health agents deliver leaflets to the older adults on their monthly home visits, with information similar to the posters that will be placed in the UBSs;

c. A website will be created (which will be informed on the poster and leaflet) with information about the study similar to the posters. This website will be accessible to the older adults and their families.

Besides the actions described above, we also contacted the Brazilian health operator Prevent Senior to discuss the possibility of including older adults who use this health plan in the two clinical trials (PRODIGITAL and PRODIGITAL-D). We are still awaiting a response from Prevent Senior on the implementation of this collaboration.

If it is possible to recruit older users of Prevent Senior, the objectives and methodology of the study will not change, only the following procedures will be added to the project:

a. Place, participants, inclusion and exclusion criteria: Older adults registered with 24 UBSs of the municipality of Guarulhos and who use the Prevent Senior health plan. We will use a list of names of the older adults provided by the UBS and Prevent Senior to perform the recruitment of the older adults;

b. Process evaluation: We will extract routine data from the health systems (UBSs and Prevent Senior) of the older adults recruited for the clinical trial;

c. Intervention and control group: The research team will not interfere in any way in the care that the older adults in the two clinical trial groups receive, regardless of the 'site' (UBS or Prevent Senior) where recruitment took place;

d. Risk assessment: We will contact the health service that the older individual uses (UBS or Prevent Senior) in situations where the participant presents 'Immediate Suicide Risk' at baseline or follow-up assessments;

e. Benefits: We believe that the inclusion of older users of UBS and Prevent Senior health plan will make the results of the study more generalizable to the Brazilian aged population;

f. The TCLEs will be the same, regardless of the place of recruitment (UBS or Prevent Senior). The wording of the TCLEs has already been adapted to be used with older people using different health services.

**TIMELINE - PRODIGITAL***

| **Month*** | **1-2** | | **3-4** | **5-6** | **7-8** | **9-10** | **11-12** | **13-14** | **15-16** | **17-18** | **19-20** | **21-24** |
| --- | --- | --- | --- | --- | --- | --- | --- | --- | --- | --- | --- | --- |
| Establishing the infrastructure |  | |  |  |  |  |  |  |  |  |  |  |
| Recruitment and training of researchers |  | |  |  |  |  |  |  |  |  |  |  |
| Contact with the Health Secretariat and UBSs of Guarulhos |  | |  |  |  |  |  |  |  |  |  |  |
| Final development of the intervention (messages) |  | |  |  |  |  |  |  |  |  |  |  |
| Ethics Committee Approval |  | |  |  |  |  |  |  |  |  |  |  |
| Clinical Trial Registration |  | |  |  |  |  |  |  |  |  |  |  |
| Pilot study |  |  |  |  |  |  |  |  |  |  |  |  |
| **Recruitment (T0):** |  | | | | | | | | | | | |
| Study server operation |  | |  |  |  |  |  |  |  |  |  |  |
| Start of recruitment (screening and baseline assessment) |  | |  |  |  |  |  |  |  |  |  |  |
| Evaluation on the acceptance of remote intervention |  | |  |  |  |  |  |  |  |  |  |  |
| **Intervention:** |  | | | | | | | | | | | |
| Intervention Group: Inform UBS, Messages (WhatsApp) |  | |  |  |  |  |  |  |  |  |  |  |
| Control Group: Inform UBS, message (WhatsApp) |  | |  |  |  |  |  |  |  |  |  |  |
| **Follow-up assessment (T1 and T2):** |  | | | | | | | | | | | |
| Start of the first follow-up assessment (T1) |  | |  |  |  |  |  |  |  |  |  |  |
| Start of the first follow-up assessment (T2) |  | |  |  |  |  |  |  |  |  |  |  |
| Data analysis, final project report, preparation of scientific papers, dissemination |  | |  |  |  |  |  |  |  |  |  |  |

*1^st^ month: August/2021; 24^th^ month: July/2023

The project schedule will be only executed after approval by the CEP.

**References**

1. Cuijpers P, van Straten A, Smit F, Mihalopoulos C, Beekman A. Preventing the onset of depressive disorders: a meta-analytic review of psychological interventions. Am J Psychiatry. 2008 Oct;165(10):1272-80.
2. Buntrock C, Ebert DD, Lehr D, Smit F, Riper H, Berking M, Cuijpers P. Effect of a Web-Based Guided Self-help Intervention for Prevention of Major Depression in Adults With Subthreshold Depression: A Randomized Clinical Trial. JAMA. 2016 May 3;315(17):1854-63.
3. Gilbody S, Lewis H, Adamson J, Atherton K, Bailey D, Birtwistle J, Bosanquet K, Clare E, Delgadillo J, Ekers D, Foster D, Gabe R, Gascoyne S, Haley L, Hamilton J, Hargate R, Hewitt C, Holmes J, Keding A, Lilley-Kelly A, Meer S, Mitchell N, Overend K, Pasterfield M, Pervin J, Richards DA, Spilsbury K, Traviss-Turner G, Trépel D, Woodhouse R, Ziegler F, McMillan D. Effect of Collaborative Care vs Usual Care on Depressive Symptoms in Older Adults With Subthreshold Depression: The CASPER Randomized Clinical Trial. JAMA. 2017 Feb 21;317(7):728-737.
4. Vaughan L, Corbin AL, Goveas JS. Depression and frailty in later life: a systematic review. Clin Interv Aging. 2015 Dec 15;10:1947-58.
5. Makizako H, Shimada H, Doi T, Yoshida D, Anan Y, Tsutsumimoto K, Uemura K, Liu-Ambrose T, Park H, Lee S, Suzuki T. Physical frailty predicts incident depressive symptoms in elderly people: prospective findings from the Obu Study of Health Promotion for the Elderly. J Am Med Dir Assoc. 2015 Mar;16(3):194-9.
6. Collard RM, Comijs HC, Naarding P, Penninx BW, Milaneschi Y, Ferrucci L, Oude Voshaar RC. Frailty as a predictor of the incidence and course of depressed mood. J Am Med Dir Assoc. 2015 Jun 1;16(6):509-14.
7. Murray E, Burns J, See TS, Lai R, Nazareth I. Interactive Health Communication Applications for people with chronic disease. Cochrane Database Syst Rev. 2005 Oct 19;(4):CD004274.
8. Johnson D, Deterding S, Kuhn KA, Staneva A, Stoyanov S, Hides L. Gamification for health and wellbeing: A systematic review of the literature. Internet Interv. 2016 Nov 2;6:89-106.
9. Gilbody S, Richards D, Barkham M. Diagnosing depression in primary care using self-completed instruments: UK validation of PHQ-9 and CORE-OM. Br J Gen Pract. 2007 Aug;57(541):650-2.
10. Löwe B, Unützer J, Callahan CM, Perkins AJ, Kroenke K. Monitoring depression treatment outcomes with the patient health questionnaire-9. Med Care. 2004 Dec;42(12):1194-201.
11. Kroenke K, Spitzer RL, Williams JB. The PHQ-9: validity of a brief depression severity measure. J Gen Intern Med. 2001 Sep;16(9):606-13.
12. Santos IS, Tavares BF, Munhoz TN, Almeida LS, Silva NT, Tams BD, Patella AM, Matijasevich A. Sensibilidade e especificidade do Patient Health Questionnaire-9 (PHQ-9) entre adultos da população geral [Sensitivity and specificity of the Patient Health Questionnaire-9 (PHQ-9) among adults from the general population]. Cad Saude Publica. 2013 Aug;29(8):1533-43. Portuguese.
13. Ell K, Unützer J, Aranda M, Gibbs NE, Lee PJ, Xie B. Managing depression in home health care: a randomized clinical trial. Home Health Care Serv Q. 2007;26(3):81-104.
14. Williams A et al. The present state of play about QALYs. In: Hopkins A, editor. Measures of the quality of life and the uses to which such measures may be put. London: RCP Publications; 1992.
15. Herdman M, Gudex C, Lloyd A, Janssen M, Kind P, Parkin D, Bonsel G, Badia X. Development and preliminary testing of the new five-level version of EQ-5D (EQ-5D-5L). Qual Life Res. 2011 Dec;20(10):1727-36.
16. Grewal I, Lewis J, Flynn T, Brown J, Bond J, Coast J. Developing attributes for a generic quality of life measure for older people: preferences or capabilities? Soc Sci Med. 2006 Apr;62(8):1891-901.
17. Sapin C, Fantino B, Nowicki ML, Kind P. Usefulness of EQ-5D in assessing health status in primary care patients with major depressive disorder. Health Qual Life Outcomes. 2004 May 5;2:20.
18. Viegas Andrade M, Noronha K, Kind P, Maia AC, Miranda de Menezes R, De Barros Reis C, Nepomuceno Souza M, Martins D, Gomes L, Nichele D, Calazans J, Mascarenhas T, Carvalho L, Lins C. Societal Preferences for EQ-5D Health States from a Brazilian Population Survey. Value Health Reg Issues. 2013 Dec;2(3):405-412.
19. Coast J, Flynn T, Sutton E, Al-Janabi H, Vosper J, Lavender S, Louviere J, Peters T. Investigating Choice Experiments for Preferences of Older People (ICEPOP): evaluative spaces in health economics. J Health Serv Res Policy. 2008 Oct;13 Suppl 3:31-7.
20. Spitzer RL, Kroenke K, Williams JB, Löwe B. A brief measure for assessing generalized anxiety disorder: the GAD-7. Arch Intern Med. 2006 May 22;166(10):1092-7.
21. Kuznier T, Oliveira F, Mata L, Chianca T. Tradução e adaptação transcultura da UCLA loneliness scale (version 3) para idosos no Brasil. Rev Min Enferm. 2016;20:1-8.
22. Kiosses DN, Leon AC, Areán PA. Psychosocial interventions for late-life major depression: evidence-based treatments, predictors of treatment outcomes, and moderators of treatment effects. Psychiatr Clin North Am. 2011 Jun;34(2):377-401, viii.
23. Schulz KF, Altman DG, Moher D; CONSORT Group. CONSORT 2010 statement: updated guidelines for reporting parallel group randomised trials. PLoS Med. 2010 Mar 24;7(3):e1000251.
24. Frederick JT, Steinman LE, Prohaska T, Satariano WA, Bruce M, Bryant L, Ciechanowski P, Devellis B, Leith K, Leyden KM, Sharkey J, Simon GE, Wilson N, Unützer J, Snowden M; Late Life Depression Special Interest Project Panelists. Community-based treatment of late life depression an expert panel-informed literature review. Am J Prev Med. 2007 Sep;33(3):222-49.
