## Supplementary material for "Self-help digital psychosocial intervention for older adults with subthreshold depressive symptoms in primary care in Brazil (PRODIGITAL): Protocol for an individually randomised controlled trial": Viva Vida programme

| **Week** | **Main topics** | **Period** | **Monday** | **Wednesday** | **Friday** | **Saturday** |
| --- | --- | --- | --- | --- | --- | --- |
| 1 | - Introduction to the programme - Psychoeducation on subthreshold depression - Introduction to the vicious and virtuous cycle of depression | M | Message 1 (audio) | Message 3 (audio) | Message 5 (audio) | Message 7 (audio) |
|  |  | A | Message 2 (image) | Message 4 (image) | Message 6 (image) | Message 8 (image)  Quick Reply message 1 |
| 2 | - Behavioural activation | M | Message 9 (audio) | Message 11 (audio)* | Message 13 (audio) | Message 15 (audio) |
|  |  | A | Message 10 (image) | Message 12 (image) | Message 14 (image) | Message 16 (image)  Quick Reply message 2 |
| 3 | - Planning activities | M | Message 17 (audio) | Message 19 (audio) | Message 21 (audio) | Message 23 (audio) |
|  |  | A | Message 18 (image) | Message 20 (image) | Message 22 (image) | Message 24 (image)  Quick Reply message 3 |
| 4 | - Review of behavioural activation - General health education | M | Message 25 (audio) | Message 27 (audio) | Message 29 (audio) | Message 31 (audio) |
|  |  | A | Message 26 (image) | Message 28 (image) | Message 30 (image) | Message 32 (image)  Quick Reply message 4 |
| 5 | - Review of psychoeducation - Review of behavioural activation | M | Message 33 (audio) | Message 35 (audio) | Message 37 (audio) | Message 39 (audio) |
|  |  | A | Message 34 (image) | Message 36 (image) | Message 38 (image) | Message 40 (image)  Quick Reply message 5 |
| 6 | - Review of planning activities - Psychoeducation on relapse prevention | M | Message 41 (audio) | Message 43 (audio) | Message 45 (audio) | Message 47 (audio) |
|  |  | A | Message 42 (image) | Message 44 (image) | Message 46 (image) | Message 48 (image)  Quick Reply message 6 |

* An extract from this audio is transcribed in the article. M: morning; A: afternoon.

Week 1: Characters welcome participants and provide information about the programme, including the number of messages, how they will be delivered, and who to contact in case of technical problems. Additionally, the characters introduce themselves, explain that they will be sharing their personal experiences of taking part in the programme, and advise participants to seek medical help if depressive symptoms persist. They describe the main signs of subthreshold depression and the mechanisms of the vicious and virtuous cycle of depression, using the metaphor of the "wheel of improvement" and the "wheel of worsening".

Week 2: Characters share how they left the vicious cycle of depression and entered the virtuous cycle of depression by starting to do activities they like and value. They encourage participants to do the same. They also talk about how these activities had a positive impact on their mood.

Week 3: Characters share strategies that participants can use to enter the virtuous cycle of depression, such as making a list of things they like to do and leaving it next to their bed, breaking activities into smaller tasks and doing them gradually, and doing activities similar to those they used to like but can no longer do. They encourage participants to use these strategies in order to facilitate behaviour change.

Week 4: Characters encourage participants to start an activity they enjoy. They also share strategies for improving sleep quality, eating healthier and being more physically active.

Week 5: Characters recall the main signs of subthreshold depression and encourage participants to make an effort to change behaviour by starting to do an activity they enjoy in order to enter the virtuous cycle of depression.

Week 6: Characters remind participants of strategies to help them plan their activities, such as breaking them down into smaller tasks and doing activities similar to those they are no longer able to do. They encourage participants to stay in the virtuous cycle of depression after the programme ends, and the importance of early recognition of recurrent depressive symptoms. Characters congratulate participants for coming this far and advise them to seek medical care if depressive symptoms persist.

In each week, the characters encourage participants to continue listening to the messages, respond to the Quick Reply messages and send audio or text messages about their experiences with the programme.
